# Dimensions and modulators of behavioural and mental-health change during the Covid-19 pandemic

**DOI:** 10.1101/2020.06.18.20134635

**Authors:** Adam Hampshire, Peter Hellyer, Eyal Soreq, Mitul A. Mehta, Konstantinos Ioannidis, William Trender, Jon E. Grant, Samuel R. Chamberlain

**Affiliations:** Imperial College London; King’s College London; Cambridgeshire and Peterborough NHS Foundation Trust; Department of Psychiatry, University of Cambridge, UK; Department of Psychiatry, University of Chicago, USA

**Keywords:** pandemic, coronavirus, Covid-19, mental health, wellbeing, mood, impulsive, compulsive, predictors, population

## Abstract

How has the Covid-19 pandemic affected mental health? What are the most common positives and negatives? How do population variables mediate the impact on mood and behaviour? Who is most at risk of adverse consequences? Which pragmatic measures can help? We address these questions in a data-driven manner by applying multivariate, machine-learning and natural-language processing methods to a survey database collected from 376,987 members of the general public. We report that small average changes in mood from pre-to mid-pandemic obfuscate substantial consequences, both positive and negative, for people from particular sub-populations, vocations, circumstances and personality profiles. The coping strategies that people find helpful during the pandemic are correspondingly diverse yet predictable. We propose that by combining psychological, and demographic variables, it is possible to identify individuals who are at most risk of adverse consequences and to extract individually tailored advice from the collective lived experiences of the general population.

## Introduction

The coronavirus disease 2019 (Covid-19) pandemic has brought about unprecedented change in peoples’ lives due to direct and indirect consequences of the illness, physical distancing and socio-economic restructuring. These changes are likely to have affected mood, anxiety, other aspects of mental health and behaviour in widespread, profound but idiosyncratic ways.^1^ Expert groups have posited that the impact on well-being is likely to relate to a variety of factors including (i) aspects of demographics such as age or ethnicity, (ii) social networks, (iii) financial/occupational circumstances (iv), being shielded or having carer responsibilities, (v) pre-existing mental health symptoms, (vi) maladaptive online technology use, (vii) personality traits and (viii) tendency towards compulsive behaviours.^2-4^

Urgent calls have been made to study these relationships because they are critical to inform policy and healthcare decisions, and to guide researchers and clinicians; however, to date, there is little published information. Indeed, knowledge about how pandemics affect mental health (including Covid-19) is limited, with many studies focusing on small rarefied samples, not examining temporal dynamics of change pre-to post-lockdown,^5^ nor integrating the diverse psycho-socio-economic variables that are relevant.^6^

More generally, addressing these knowledge gaps during the pandemic provides a unique opportunity to understand the nature and psychological basis of mental health resilience and vulnerability in the general population. It also presents a major methodological-statistical challenge. Specifically, there is likely to be a many-to-many mapping relationship between the ways that people have been affected by the pandemic and their psycho-socio-economic profiles. Furthermore, many of the relevant variables, for example psychiatric traits and technology use behaviours, will tend to covary. Moreover, it is not altogether clear what the major dimensions of the impact are, or which coping measures can help. In such a context, identifying, disentangling and mapping the key variables can only be achieved in a data-driven multivariate manner, which in turn necessitates the collection and analysis of largescale population data.

To address this challenge, we applied a combination of multivariate, machine learning, and natural-language processing methods to analyse a unique large-scale dataset, comprising a survey of mental health and wellbeing variables completed by 376,987 people, collected since late December 2019 and focused on January and May 2020, in collaboration with BBC2 Horizon, predominantly within the UK. Within this broader database were responses to a comprehensive questionnaire instrument capturing self-reported pandemic-impact in 74,830 out of 110,118 respondents in May, and free text from ∼50,000 respondents stating in their own words the main positive and negative consequences of the pandemic, and what they had found helpful and would recommend to others to maintain health and well-being during this time.

We first sought to confirm whether there were global differences in the population distributions of depression, anxiety, and sleep problems between January, just prior to the Covid-19 outbreak reaching the UK, and May, during peak lockdown. Next, we estimated in a data-driven manner the dimensionality of self-perceived impact of the pandemic during May. We then identified the major predictors for each impact dimension, in terms of sociodemographic, economic and individual-circumstance variables (e.g., work and home arrangements), pre-existing mental and neurologic disorders, personality and psychiatric traits, and technology use. Finally, we analysed free text data to determine the diversity of strategies that people most commonly reported helped them to cope during the pandemic, characterise those topics, and then explore the potential to generate individually tailored advice by mapping the covariance between topic prevalence and the population variables in a multivariate manner.

## Results

### Respondents

Starting from December 26th 2019, participants were recruited to the study website, where they completed cognitive tests and a detailed questionnaire (**Supplement 1**). Articles describing the study appeared on the BBC2 Horizon page, BBC Home page, BBC News Home page and circulated on mobile news meta-apps from January 1^st^ 2020. A second promotional drive was launched on May 2^nd^ 2020, aligned with a BBC2 Horizon Documentary focused on concurrently collected cognitive data, analysis of which is outside the scope of the current article. This produced two large peaks in data collection after the January (pre-pandemic N= 225,437) and May (mid-pandemic N= 122,680) launches, with more minimal data (Early-pandemic 9,610) in the intervening period. Participants aged under 16 were excluded prior to analysis because they were presented with an abbreviated questionnaire. Also, as a quality control people who completed the questionnaire too rapidly (defined as under 6 minutes) were excluded. This resulted in 215,886 Pre-pandemic, 8,680 Early-pandemic and 110,118 Mid-pandemic datasets in the reported analyses (**Figure 1a**).

**Figure 1.**
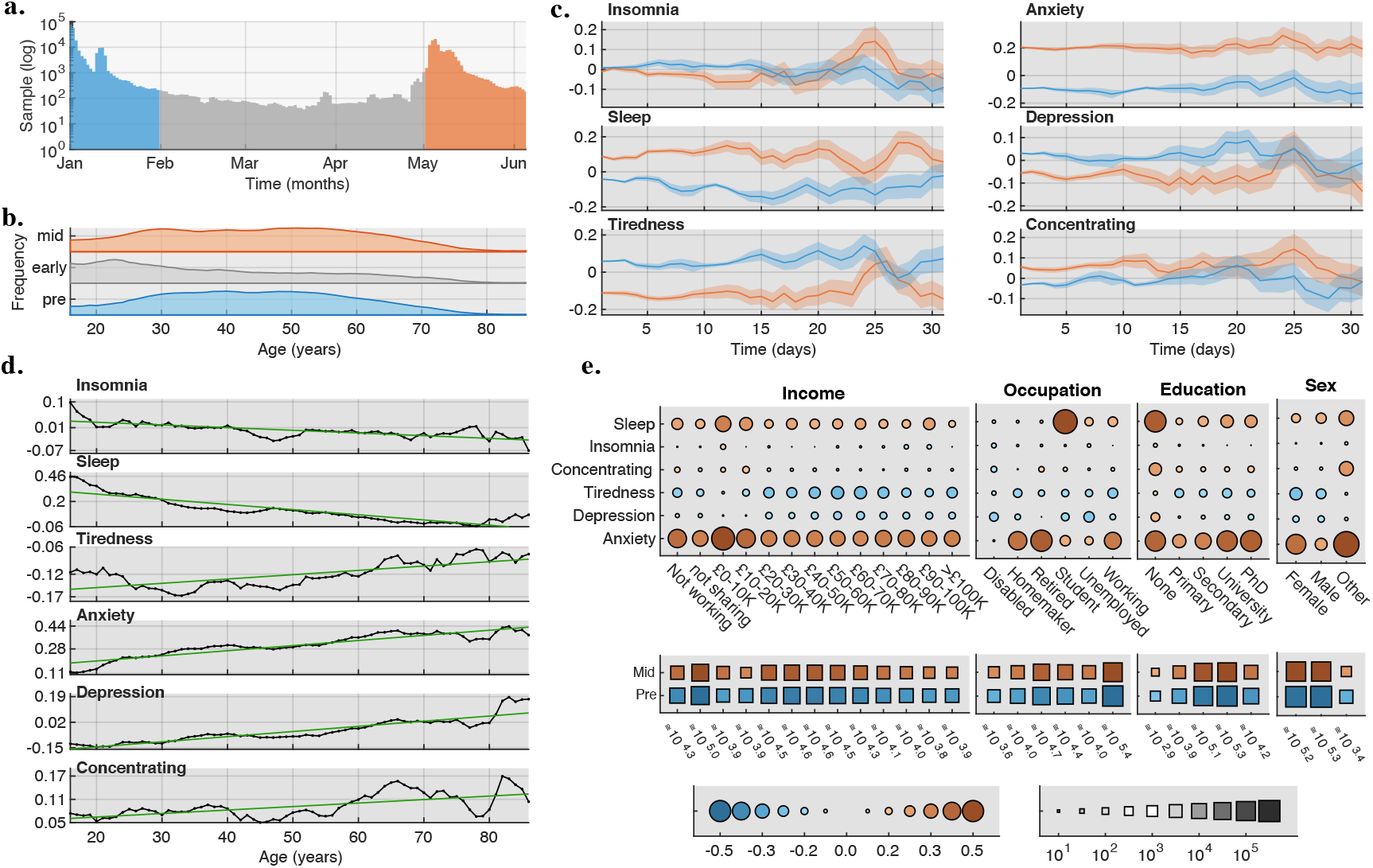
Sociodemographic mediated differences in national mood scores during the Pre- and Mid-Pandemic epochs. **1a.** Data were collected throughout the first 5 months of 2020. Sampling was concentrated in early January and early May, when BBC2 Horizon, News and Homepage promoted the study. 1b. Sociodemographic distributions, including age, were closely matched Pre-Pandemic (January) and Mid-Pandemic (May). Early-Pandemic (February-April) was more sparsely sampled and captured people from a different age distribution. 1c. Daily mean with SEM mood scores of the population, calculated separately for each of the 31 days post promotion launches Mid-Pandemic and Pre-Pandemic. Age, gender, handedness, first language, country of residence occupational status, and earnings are factored out. Significantly scaled differences are reproducibly evident across the days, most notably increased prevalence of mood-anxiety, but also improved sleep and tiredness scores. 1d. Differences in mean mood scores Mid-Pandemic minus Pre-Pandemic related to age. Older adults had a greater increase in anxiety. Younger adults increased sleep. Younger adults were less depressed whereas older adults were more depressed. 1e. Sub-population counts (middle squares - size represents log N per sub population) and the corresponding scale/valence of mood-score change (upper circles - size represents SD units and colour direction of change). Substantial differences were evident as a function of sociodemographic sub-groups, with heightened anxiety particularly in retired people, workers, homemakers, low income earners, and for gender Other vs Female vs Male.

Plotting sociodemographic distributions for each of the three epochs (**Figure 1b&e**) showed high correspondence in sociodemographic distributions including sex, handedness, first language, ethnicity and education level. However, the more sparsely sampled early-pandemic epoch age distribution had a marked skew towards younger age, likely a consequence of reduced visibility within the public eye at that time. The other two epochs captured a broad and inclusive cross section of the population, with >100 participants per age year up to and including 85. Participants above this age were combined into an 86+ category. ∼15% indicated that they were from minority ethnic groups. There were 161,916 female, 187,622 male and 3,026 indicating other (i.e. non-binary) gender. This demonstrated appropriate representation of minority groups in the dataset.

### Differences in the population distributions of mood scores Pre-vs Mid-Pandemic

Differences in mood assessment scores were calculated for the densely sampled and closely matched Mid vs Pre-Pandemic epochs after factoring out age, sex, handedness, education level, first language, country of residence, occupational status and income. Six mental health measures were calculated and compared separately for people who were sampled at each of 31 days post launch, effectively providing 31 independent replication analyses (**Figure 1c)**. When analysing large-scale data, very small effects will tend to have highly significant statistical values. Therefore, the better measure of significance is effect size. Here, the mean differences in mental health measures were generally in the small effect size range. The most reliable difference during the Mid-pandemic vs. Pre-pandemic epochs was for anxiety, the incidence of which increased on average by ∼0.3 standard deviation units (SDs), with this result reliably evident in all 31 analyses. Depression decreased on average by ∼0.08 SDs, an effect that was only evident in the more densely sampled timeframe just post launch. A decrease in tiredness was evident for almost all days, with an average difference of ∼0.16SDs. Problems concentrating increased marginally ∼0.07SDs. Problems with insomnia stayed approximately the same; however, mean reported hours slept per night increased ∼0.19SDs.

General Linear Models (GLMs) with interaction terms showed that these small but robust overall differences in mental health measures were amplified in select sociodemographic sub-populations (**Figure 1d-e**). Older adults showed the greatest increase in anxiety incidence, (0.4-0.5SD for 60-80-year-olds). Increases in depression was lower for younger adults and higher for older adults (−0.2SDs to 0.1SDs). Females (0.32SDs) had a greater increase in anxiety than Males (0.21SDs) but less than those identifying their gender as Other (0.38SDs). Vocationally, increased anxiety incidence was greater for retired people (0.38SDs), followed by homemakers (0.32SDs) and workers (0.29SDs). People who identified gender as Other reported the greatest increase in problems concentrating (0.22SDs).

### Self-perceived pandemic impact and the PD-GIS-11 scale

The results of the cross-epoch analyses provided initial evidence that there might be disproportionate pandemic effects on the above-noted aspects of mental health for some people. A limitation though was that any finer grained behavioural basis to the pandemic impact could not be elucidated from the mood questionnaire alone. Consequently, in May, the online questionnaire was extended with the PanDemic General Impact Scale (PD-GIS-11), designed to probe self-perceived impact of the pandemic on day-to-day life.

Analysis of responses to individual items of the Pandemic General Impact Scale (PD-GIS-11) for 74,830 people showed strong overall agree/disagree biases for a subset of positive and negative statements (**Figure 2)**. Some of the strongest agreement was with positive statements, e.g., improved natural environment, enjoying the simpler things in life, spending less money, saving more money, and a greater sense of community. Amongst the strongest agreement with negative statements was concern for health of loved ones, which notably, was higher than concern about one’s own health, and loss of leisure/health activities. The strongest disagreement was with statements regarding loss of employment, increased conflict at home, preoccupation with infection and loss of access to basics. There was strong agreement that technology/science/healthcare would advance more rapidly and that things would change but not necessarily for the worse, but strong disagreement that economic impact would be temporary. People agreed that communication apps helped stay in touch with loved ones. These measures of overall agreement were contextualised by substantial population variability. Together, these results indicate that people were affected in very different ways, and not all of them negative.

**Figure 2.**
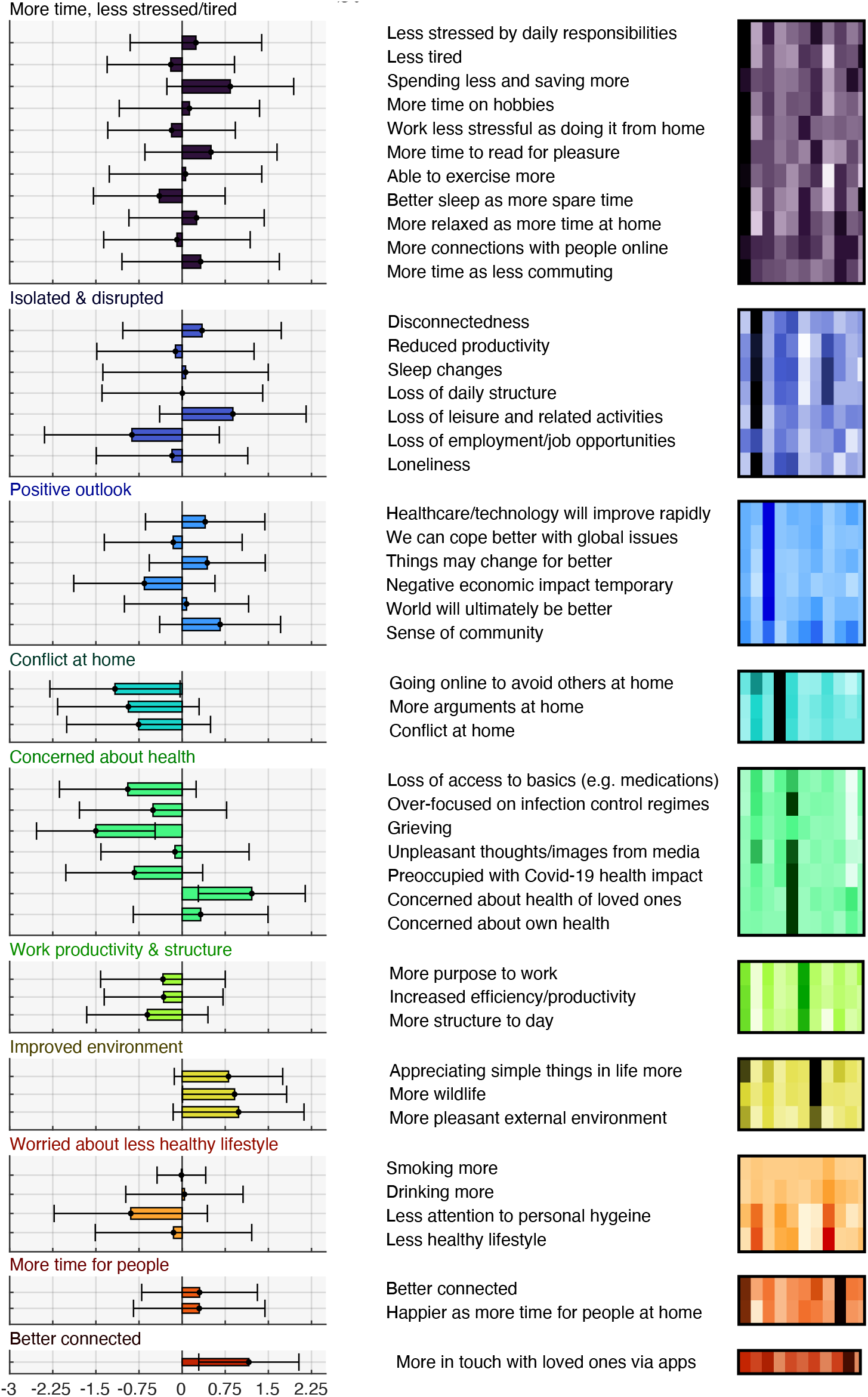
Individual item analysis and principal component analysis for the PD-GIS-11. **Left.** Strength of agreement with statements about pandemic impact. X scale is in SD units. **Right**. PCA identified 11 components underlying PD-GIS-11 responses (**Supplement 3**). These were labelled 1: positive perception of more free time, less stress and reduced tiredness. 2: loneliness/ disruption. 3: positive outlook. 4: increased conflict at home. 5: increased health obsession. 6: increased work engagement/efficiency. 7: improved environment. 8: concern about unhealthier lifestyle. 9: time for people. 10: connectedness. 11 Better sleep.

Principal component analysis (**Supplement 2**) was conducted on the PD-GIS-11 with varimax rotation. Application of the Kaiser convention of including components with eigenvalues > 1 indicated 11 orthogonal dimensions, further supporting the view that people were affected in different ways. The component capturing the most variance (1) had heavy loadings for questions pertaining to positive perception of more free time, less stress and reduced tiredness. The next largest component (2) represented questions pertaining to loneliness and disruption of normal life. Subsequent factors represented (3) positive outlook; (4) increased conflict at home; (5) increased health obsession; (6) increased work engagement/efficiency; (7) improved environment; (8) concern about unhealthier lifestyle; (9) time for people; (10) connectedness; and (11) better sleep (**Figure 2**).

Bivariate cross-correlation of the PD-GIS-11 component scores with the mood self-assessment items demonstrated that self-perceived impact of the pandemic explained a substantial component of variance in the incidence of negative mood symptoms (**Figure 3**). This was primarily the case for the negative-impact sub-scales of the PD-GIS-11. The robustness of this relationship was further corroborated via canonical correlation analysis, which produced 10 statistically significant correlation modes, the largest of which had a canonical r value of 0.62 (p<0.001).

**Figure 3.**
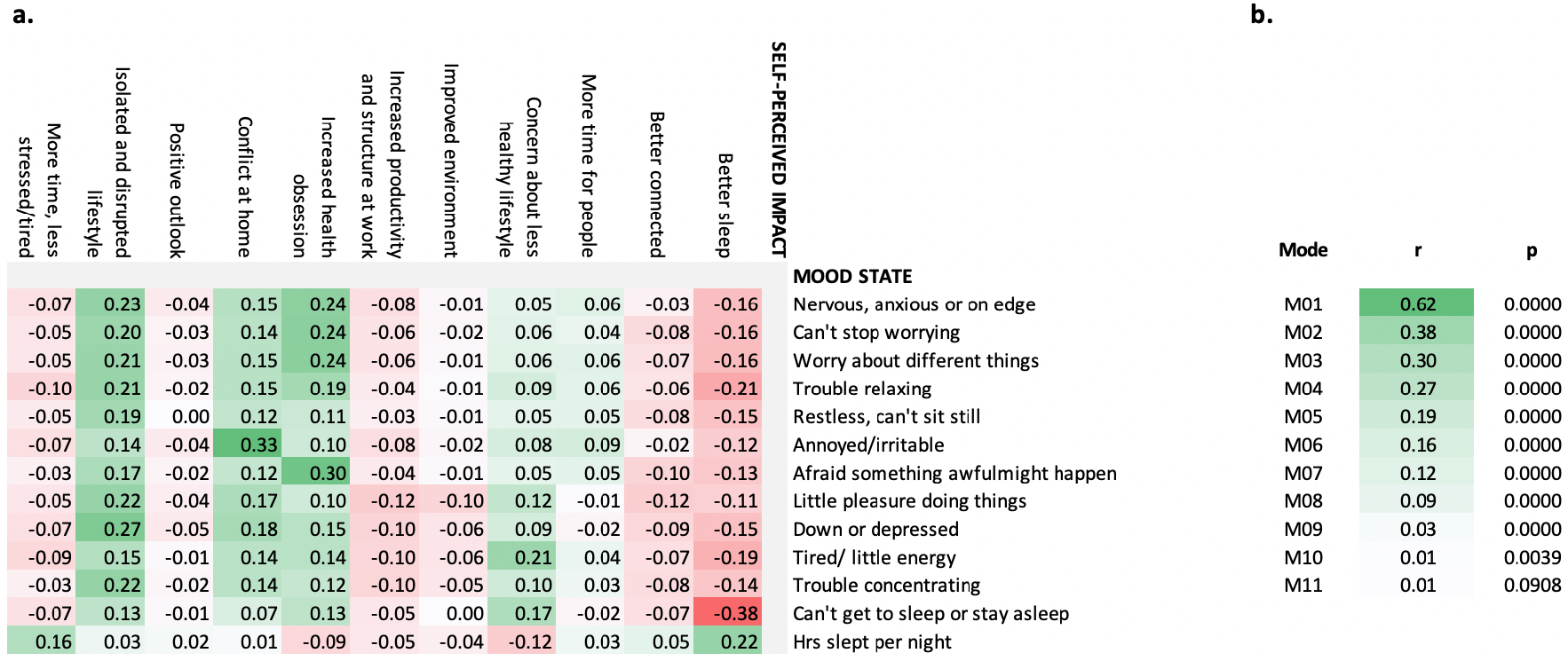
Interrelationship between the PD-GIS-11 sub-scales and mood assessment. **a.** Pearson’s r values for questionnaire scale pairwise correlations. Significant correlations were evident between measures of self-perceived covid-19 pandemic impact, as captured in the PD-GIS-11 sub-scales, and the mood self-assessment items. These were clearly stronger for the negative impact sub-scales. **b.** Canonical correlation analysis confirmed this relationship in a multivariate manner, with 10 statistically significant correlation modes, the largest of which had a canonical r value of 0.62.

### Statistical predictors of self-perceived pandemic impact

Together the above results demonstrated the validity of the PD-GIS-11 scale in providing a multi-dimensional assay of the idiosyncratic ways in which the pandemic affected mood and behaviour in May, during the lockdown. This enabled the central question, regarding how population variables relate to pandemic impact to be addressed.

We first used GLMs to examine the relationships of PD-GIS-11 scores with socio-demographic and economic variables, home context, cohabitees and work arrangements (**Figure 4**). A complex multivariate set of factors were substantially predictive of PD-GIS-11 scores (**see Supplement 3 for full results**). Work arrangements were amongst the most prominent with vocational differences spanning a very large effect size range; for example, health workers, particularly those on the frontline with Covid-19 patients had less time and were less relaxed relative to those who had been furloughed (1.5SDs), but also reported sleeping better (0.34SDs), having greater engagement in work (0.67SDs), and were the most likely to agree that the natural environment had changed for the better (0.25SDs).

**Figure 4.**
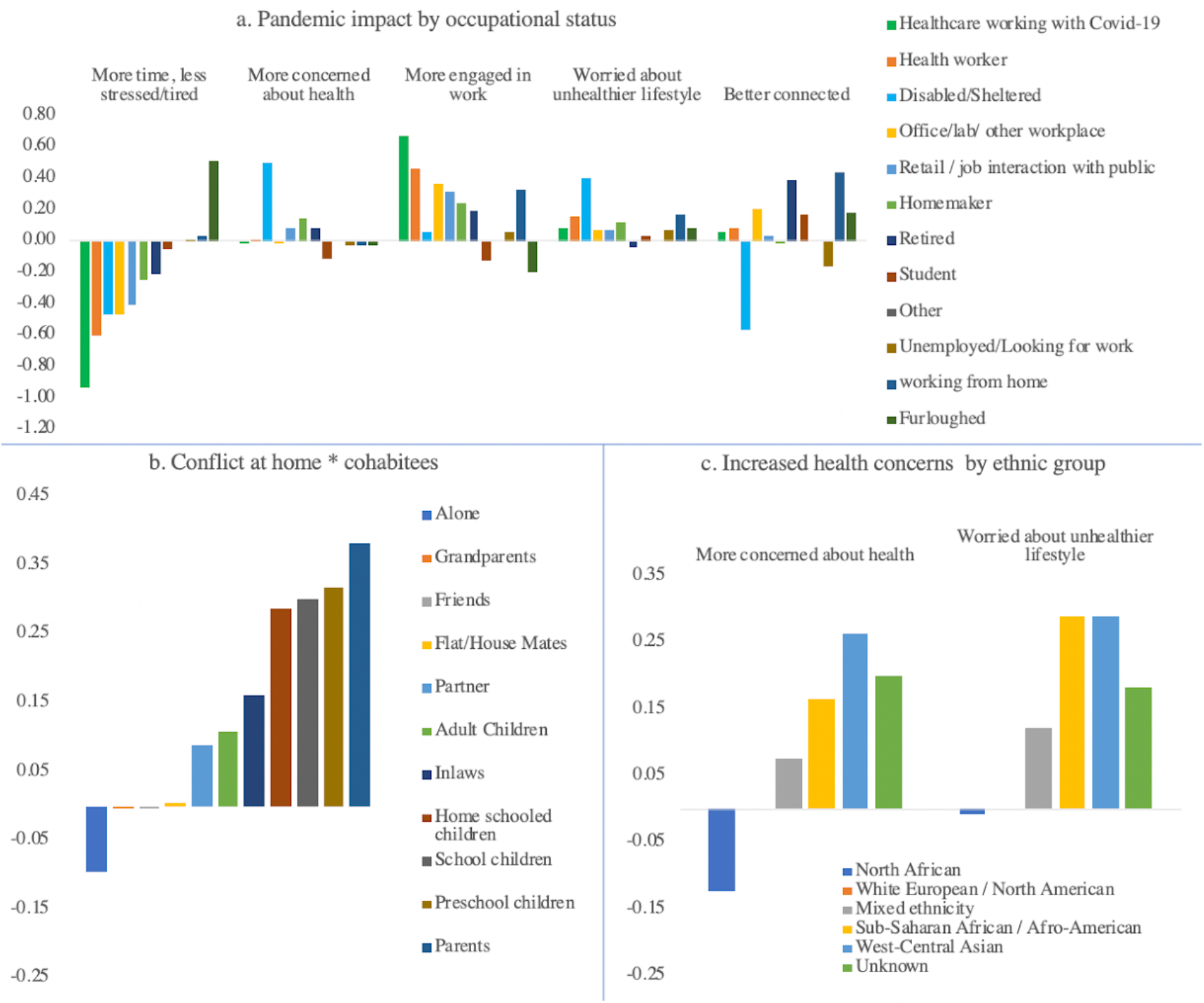
Impact of the Covid-19 pandemic as a function of selected demographic variables. **a.** There were large and disproportionate impacts of the pandemic on the wellbeing of healthcare workers and disabled/shielded people. **b.** Conflict at home was predicted by type of cohabitees. **c.** Ethnicity predicted increased health concerns. **d.** People with anxiety and obsessive-compulsive disorder (OCD) were disproportionately concerned about health. **e.** People with attentional deficit hyperactivity disorder were more likely to report increased conflict at home. All effects reported in standard deviation units. Full GLM results are in **Supplement 3**.

People whose incomes had been negatively affected reported disproportionate disruption to everyday life (0.45SDs). People identifying as disabled or shielded reported substantially higher health obsession (0.5SDs), were less likely to report better connectedness (−0.57SDs) and were amongst the least likely to report feeling less stressed and having more time (−0.46SDs). The strongest contextual predictors of increased conflict in the home were cohabiting with preschool or school children (0.32SDs & 0.3SDs) or living with parents (0.38SDs)/in-laws (0.16SDs). People who had no, or unpleasant, outside environmental space were less likely to report reduced tiredness/stress than those with relaxing outside space (0.31SDs), were more worried about negative health implications of the lockdown (0.43SDs), and were less likely to report improved natural environment (0.43SDs). People from minority ethnic backgrounds tended to report higher health concerns (0.15-0.30SDs), but also were more likely to have a positive outlook (0.05-0.27SDs). The 8,347 participants who indicated they were looking after vulnerable older adults had higher scores for health obsession (0.17SDs) than the cohort average.

Next, we examined PD-GIS-11 scores for people reporting established diagnoses of different mental health and neurological conditions (**Figure 5 & Supplement 3**) after factoring out the above sociodemographic variables. As expected, depression (10,526) and anxiety (10,525) overlapped heavily in terms of comorbidity (6,305). Relative to the population mean, individuals with anxiety disorder and/or obsessive-compulsive disorder (797) reported greater increases in health obsessions relative to those reporting no conditions (0.25SDs). Those with attention-deficit hyperactivity disorder (ADHD, 449) reported greater increase in conflict at home (0.17SDs), and individuals with depression were more worried about having unhealthier lifestyles (0.18SDs). All clinical groups, apart from people with Parkinson’s Disease, did not experience an increase in connectedness during the pandemic to the same extent as people without these disorders (0.05-0.21SDs).

**Figure 5.**
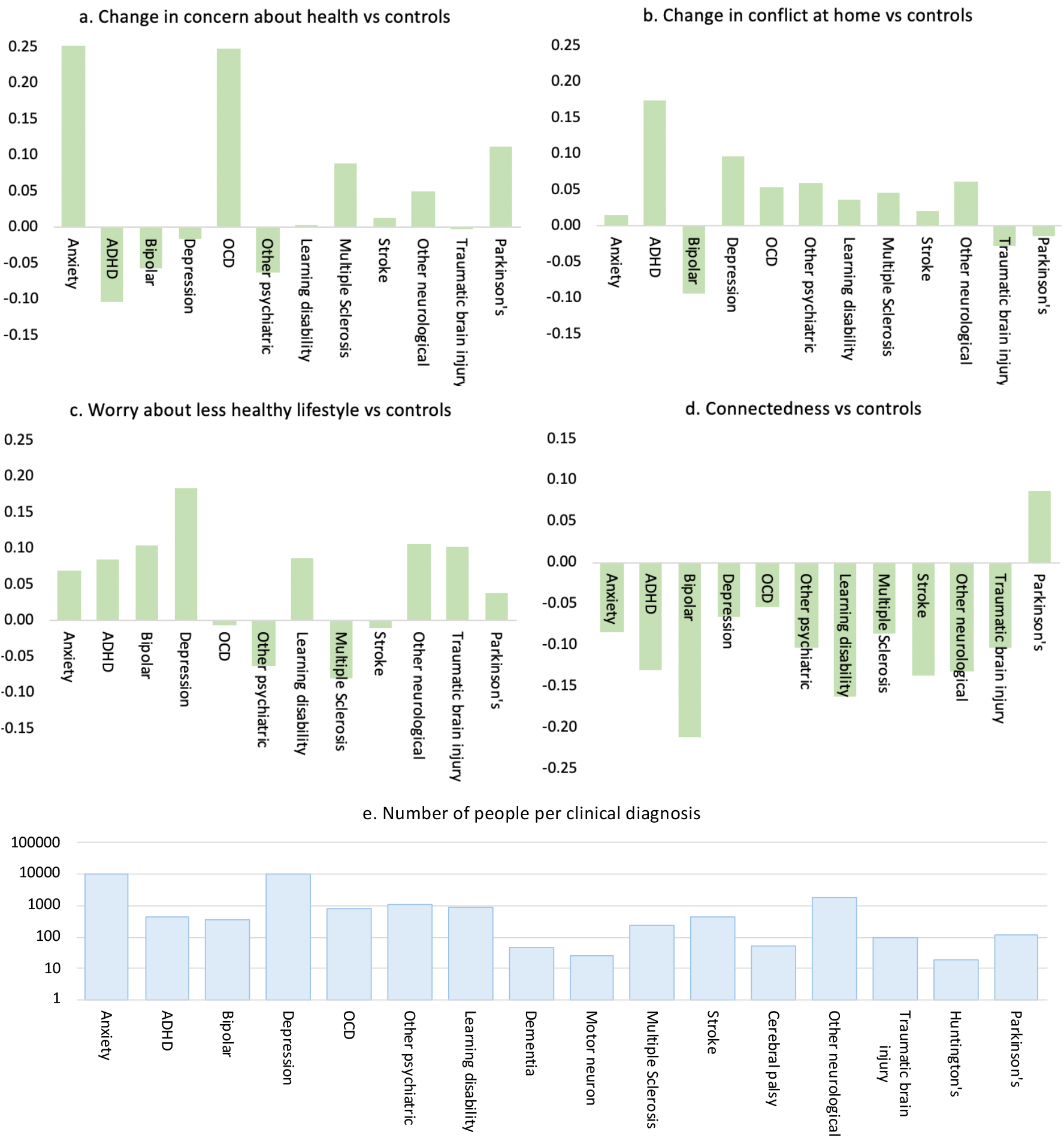
Impact of the Covid-19 pandemic as a function of neurologic and psychiatric conditions. (Y axes for effect sizes in SD units). People who had been diagnosed with pre-existing conditions were more likely to report adverse consequences of the Covid-19 pandemic. **a**. Obsessive compulsive disorder and anxiety were associated with increased concerns about health during the pandemic. **b**. Attention deficit hyperactivity disorder was associated with increased conflict in the home. **c**. People with a history of depression were more likely to report increased concerns about the impact of the pandemic on the healthiness of their lifestyle. **d**. Almost all groups showed reduced connectedness relative to controls, with the exception of people with Parkinson’s. **e**. Number of people indicating that they had been diagnosed with pre-existing neurologic and psychiatric conditions.

Finally, canonical correlation analysis was applied to quantify the multivariate associations between online technology use, personality traits, and compulsivity with Covid-19 impact (**Figure 6**) after factoring out sociodemographic variables. There were 11 statistically significant correlation modes. Full analyses are provided in **Supplement 3**. Cross-validation analysis confirmed the model was not over-fitted, which is to be expected given the very large sample size. Back projecting the first CCA mode (canonical r=0.41) onto the PD-GIS-11 showed strong associations with adverse impact, including isolation/disruption (r=0.44), increased health obsession (r=0.58) and increased conflict at home (r=0.27). Back projection on the other side of the first mode showed strong associations with technology addiction (r=0.67), stress arising from technology use (r=0.52), compulsivity subscale reward drive (r=0.45) and cognitive rigidity (r=0.38), and negative association with the personality trait ‘self security’ (r=−0.75). Back projecting the second mode (canonical r=0.34) onto the PD-GIS-11 summarised positive pandemic impact, including better connectedness (r=0.55), improved environment (r=0.39), positive outlook (r=0.35), and greater work engagement (r=0.27), with a negative relationship to conflict at home (r=−0.26). The other side of the second mode associated positively with the traits compassion (r=0.69), conscientiousness (r=−0.31) and perfectionism (r=0.32), and negatively with introversion (r=−0.28). Therefore, the self-reported impact of the pandemic was substantially mediated along multiple dimensions by personality traits, compulsivity, and technology use.

**Figure 6.**
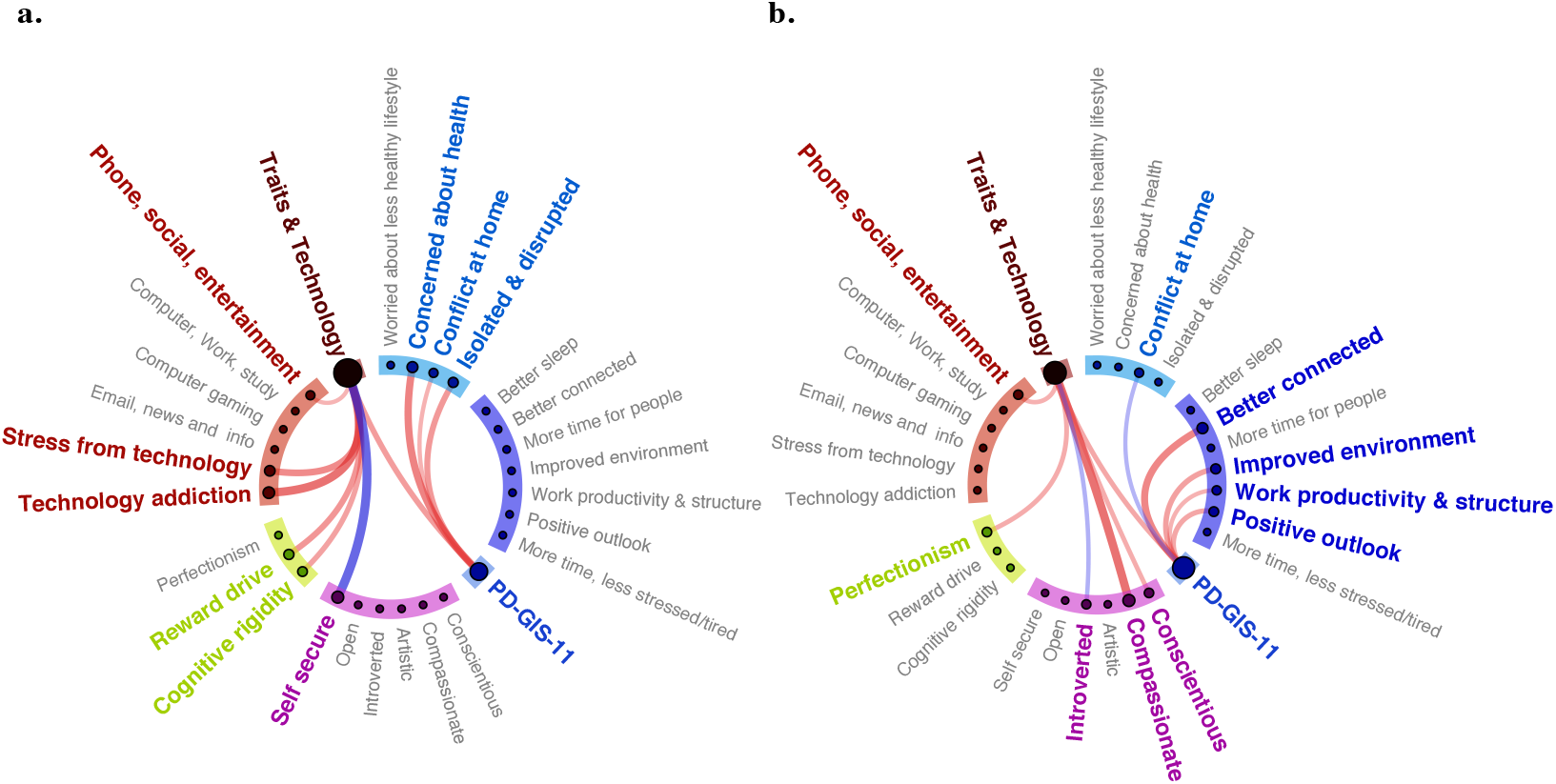
Canonical Correlation of Trait and Technology vs PD-GIS-11. Correlation weights of the trait & technology vs PD-GIS-11 canonical variates with each other and their back projections, thresholded at | r | >0.25 (full results in **Supplement 3**). **a**. Mode 1 associated negative aspects of pandemic impact with technology stress and addiction, compulsivity traits and insecurity. **b**. Mode 2 associated positive aspects of pandemic impact with positive personality traits and perfectionism, a compulsivity trait. Red lines = positive correlations. Blue lines = negative correlations. Line weight = correlation r values.

### Data-driven analysis of common impact and advice topics from free text

We further examined the dimensionality of pandemic impact through the application of Latent Dirichlet Allocation to free text in response to the questions “What has been most POSITIVE about the lockdown?”, “What has been most NEGATIVE about the lockdown?” This is one of the most established methods for identifying commonly co-occurring combinations of words, which can be used to characterise each document according to the topic mixtures from which it is comprised. This allows the prevalence of the topics to be further examined in relation to population variables of interest.

The best fitted ten exemplars of each topic from the free text are provided in **Supplement 4a**. The optimal number of topics was identified by calculating models at different levels of dimensionality in steps of five and determining the one that produced the lowest perplexity measure. Descriptions of the main negatives from 49,482 participants (**Figure 7a**) were best summarised by just five topics. In prevalence order, these were being unable to see relatives (especially older relatives), impracticalities of working or schooling from home, disruption of social and recreational activities, physical and mental health worries, and frustration with the media or government. Descriptions of the main positives from 48,315 people (**Figure 7a**) were somewhat more diverse, summarised by 10 topics, these being more free/recreational time, improved environment, time for important things, slower pace of life, positive long-term change, more time with family, learning new skills/expertise, more regular digital contact with friends/family, and enjoying the outdoors.

**Figure 7.**
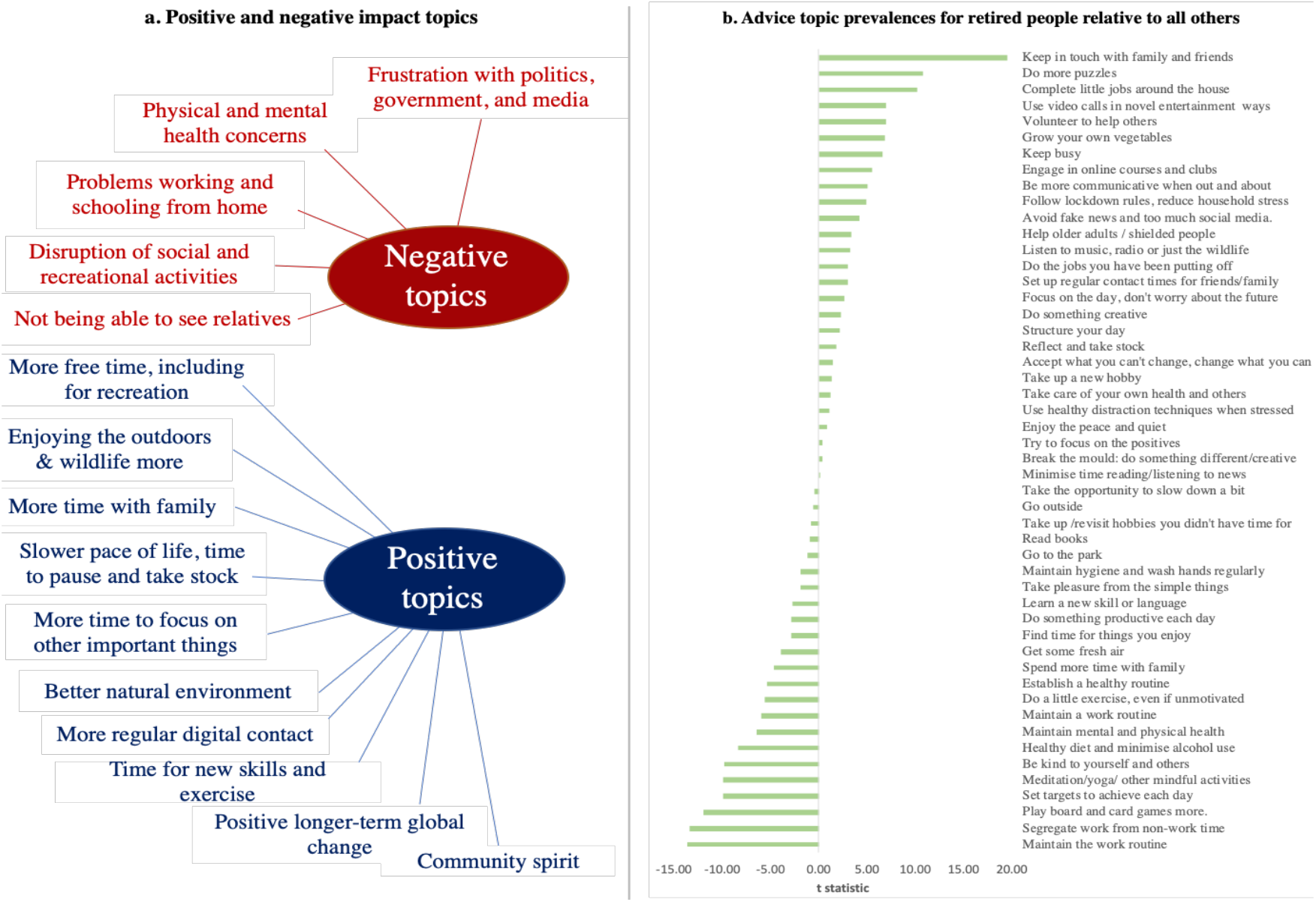
Topic modelling of free text responses. **a**. Latent Dirichlet Allocation (LDA) identified 5 negative topics and 10 positive topics from the free text responses describing the main negative and positive impacts of the Covid-19 pandemic. **b**. LDA identified ∼50 topics from free text describing what people would recommend to others based on their lived experiences of the pandemic. The prevalence of the topics varied significantly across sub populations. Above, we present differences in the advice topic prevalence for retired older adults relative to the rest of the cohort.

The same approach was then applied to free test responses to the question “What has been most POSITIVE about the lockdown?” Advice from 44,376 people was much more variable, being optimally summarised by ∼50 topics. A number of topics fitted within the broader themes of structuring time, keeping occupied, maintaining communication and keeping physically healthy. The most prevalent topic was establishing a healthy routine, followed by using video conferencing to stay in touch, meditation/yoga, regular exercise, time outside, keeping busy, going for regular walks and planning a healthy diet.

When concatenating topic prevalence estimates, there was substantial shared variance between peoples’ topic mixtures and both the mood self-assessment (**Supplement 4b**) and the PD-GIS-11 (**Supplement 4c**), as quantified using canonical correlation analysis. This provided a cross validation of the scales and free text.

A key question was whether the coping strategies that people recommended differed for people at greater risk of negative pandemic impact. To address this question, t-tests examined whether the topic prevalence covaried with some of the risk factors identified above (**Supplement 4d**). Some of the largest differences in topic prevalence related to retirement status (**Figure 7b**). Most prominently, retired people were more likely to recommend ‘set up regular time to keep in touch with friends and family’ (t=19.506 p<0.001) and ‘doing more puzzles’ (10.729 p<0.001) and taking time to complete jobs around the house (t= 10.223 p<0.001). As one would expect, work related topics were less prevalent, including ‘segregate work and non-work time (t=−13.330 p<0.001) and ‘maintain the work routine’ (t=−13.580 p<0.001).

People whose income had been negatively affected were more likely to recommend ‘go to the park regularly’ (t=5.196 p<0.001) but less likely to recommend ‘take the opportunity to slow down a bit’ (t=−7.057 p<0.001). Frontline health workers were more likely to recommend ‘maintaining personal hygiene/handwashing’ (t=3.199 p=0.001), and ‘segregating work and home time’ (t=3.102 p=0.002). Conversely ‘structure your day’ (−3.421 0.001) and ‘use distraction techniques when stressed’ (t=−3.239 p<0.001) were less relevant for them. In contrast, more prevalent topics amongst disabled and shielded people included ‘doing exercise even if unmotivated (t=3.918 p<0.001), ‘be more communicative with people’ (t=4.703, p<0.001), ‘follow lockdown rules and work out new approaches to reduce stress in the household’ (t=3.524 p<0.001), ‘listen to music or the wildlife’ (t=3.878 p<0.001) and ‘use the time to reflect’ (t=4.253 p<0.001).

People who had been diagnosed with anxiety or depression were more likely to recommend ‘being kind to yourself and others’ (anxiety t=4.996 p<0.001; depression t=7.863 p<0.001) and to ‘listen to music or appreciate wildlife’ (anxiety t=3.742 p<0.001; depression t=4.321 p<0.001). Those with anxiety also were more likely to recommend meditation or yoga (anxiety t=4.552 p<0.001; depression t=−0.278 p=0.787), whereas those with depression were more likely to recommend ‘taking pleasure in simple things’ (anxiety t=1.687 p=0.0917; depression t=3.868 p<0.001) and ‘doing exercise even if unmotivated’ (anxiety t=2.468p=0.014; depression t=3.438 p<0.001).

Finally, the prevalence of advice topics was examined in relation to people living with children because this was amongst the most substantial predictor of pandemic impact. The majority of topics also differed significantly in prevalence. Most notably, ‘do something creative’ (t=13.282 p<0.001), ‘take the opportunity to slow down a bit’ (t= 9.021 p<0.001), ‘try to focus on the positives’ (t=7.921 p<0.001) and ‘follow lockdown rules and work out new approaches to reduce stress in the household’ (t=7.254 p<0.001) were all more prevalent.

Therefore, despite the intrinsic noisiness of topic modelling, the broad range of pragmatic coping strategies that people recommend applying differed significantly for sub-populations whose were most at risk of adverse impact during the pandemic.

## Discussion

Our results provide converging evidence at large population scale that by May 2020 (peak UK lockdown), the Covid-19 pandemic had profound but idiosyncratic effects on mental health and wellbeing in the UK. The effects were complex, comprising multiple dimensions of change, some being positive. The substantial variability in these dimensions of impact was associated with a combination of clinical/biological and psycho-socio-economic variables. Furthermore, there were statistically robust relationships between these variables and what people reported, in their own words, had helped them during the pandemic.

Differences in the population distributions of depression, anxiety, and sleep were observed from pre-pandemic to the time of maximal UK lockdown, being indicative of more pronounced untoward effects in particular groups, such as in older adults. However, such macro-data obfuscates considerable nuanced variations in the nature and extent of pandemic impact across individuals. Demographic characteristics, work, environment and social circumstances all had robust associations, varying in scale from small to very large, with the nature and extent of self-reported impact of the pandemic. In some ways this was anticipated based on extant literature,^8-12^ but in others it was unexpected. Most notably, health workers showed very large differences to the broader population, being less relaxed and having less free time, but also reporting better sleep and greater work engagement. People from minority backgrounds were more worried about the impact of the pandemic on health, but also had more positive outlooks.

It was predicted that the impact of the Covid-19 pandemic would be substantially influenced by mental health and neurologic disorders,^1,2^ and dimensional traits.^13^ Notably, although overall, people with psychiatric and neurologic conditions were less likely to report increased connectedness during the pandemic, these generalised associations were of small scale. More selective associations were observed for particular disorders: e.g. elevated health worries in anxiety disorders / obsessive-compulsive disorder, and increased conflict at home in adults with attention-deficit hyperactivity disorder.^14,15^ However, trait and personality scales collectively accounted for disproportionate pandemic impact: negative impact was associated with compulsivity and insecurity, and positive impact with compassion, conscientiousness, perfectionism, and extroversion. This accords with the notion that certain personality traits are prominent in shaping resilience, whereas others engender vulnerability.^16^

Online technology can provide a powerful mediator of positive and negative impact during the pandemic. Prior studies proposed that a subset of people develop problematic usage of online technology.^3,17,18^ Accordingly, negative Covid-19 impact was strongly linked not with time spent using online technology *per se*, but with maladaptive online behaviours.

Conversely, the benefits of using technology to stay connected were prominent in the questionnaire and free text analyses. One might conclude that limiting screen time could be counterproductive; more nuanced approaches to develop healthy online behaviours are warranted.

The profound impact of the pandemic was perhaps best captured in the data-driven analysis of free text describing peoples’ lived-experience. Crucially, the dimensionality of advice was much higher than for positive and negative impact statements, with ∼50 topics optimally summarised the practical measures people said helped them during the pandemic. On one level, the fact that the prevalence of these topics differed significantly as a function of participant characteristics is surprising insofar as topic modelling is intrinsically noisy and the diversity of possible topics very broad. On another level, the observed relationships in many cases make intuitive sense; some measures are irrelevant or impractical, whereas others are more germane, depending on one’s individual circumstances. Looking forwards, we believe that there is potential to extend the preliminary topic modelling analysis conducted here in order to identify pragmatic individually tailored advice based on this novel intersection of sociodemographic, contextual and personality variables with the corpus of peoples’ self-expressed expert experiences. The rationale for thinking this could be effective as an intervention approach is simple: if something has proven tractable and effective for many people, in their own experience, it likely will also prove helpful for others. Indeed, we propose that such topics could conceivably be updated on a periodic basis, to determine pragmatic measures that people find helpful as events unfold during the recovery phase and beyond, and then making that advice available to others who have matched profiles. Future research should explore this potential through deeper analysis of the topics and study as a potential pragmatic intervention in future research.

Taken together, these results demonstrate the importance of measuring multiple dimensions when quantifying pandemic impact on mental health, and the necessity of incorporating the broad psycho-socio-economic context when seeking to understand, predict or mitigate such impact. The largest associations explaining pandemic impact related to occupation and home context, followed by medium associations with personality traits, compulsivity and maladaptive technology use, and smaller but still notable associations for mental health and neurological disorders and demographic characteristics. This complexity in turn necessitates careful study design to account for confounding factors.

Primary limitations of this study pertain to cross-sectional analysis albeit within the context of a longitudinal study. This is somewhat mitigated by the large scale of data and matching demographics for Pre and Mid pandemic timeframes, and by the rigorous multivariate statistical approaches deployed. We intend to recontact this cohort at 3, 6 and 12 months to plot change in the idiosyncratic impact of the pandemic and its aftermath in vivo^19,20^ including more detailed assessment of identified vulnerable sub-groups. We also note that the current paper focused on data from people aged 16 and above. It is vital that research also examines younger people using this and related methodologies adapted for that purpose.^2^

## Methods

### Recruitment

Starting from December 26th 2019, participants were recruited to the study website, where they completed cognitive tests and a detailed questionnaire. Articles describing the study were placed on the BBC2 Horizon, BBC Home page, BBC News Home page and circulated on mobile news meta-apps from January 1^st^ 2020. To maximise representativeness of the sample there were no inclusion/exclusion criteria. Analyses here exclude data from participants under 16 years old, as they completed a briefer questionnaire, and those who responded to the questionnaire unfeasibly fast (<4 minutes). Cognitive test data will be reported separately. The study was approved by the Imperial College Research Ethics Committee (17IC4009).

### Data collection

Data were collected via our custom server system, which produces study-specific websites (https://gbws.cognitron.co.uk) on the Amazon EC2. Questionnaires and tests were programmed in Javascript and HTML5. They were deliverable via personal computers, tablets and smartphones. The questionnaire included scales quantifying sociodemographic, lifestyle, online technology use, personality, and mental health (**Supplement 1**). Participants could enrol for longitudinal follow up, scheduled for 3, 6 and 12 months. People returning to the site outside of these timepoints were navigated to a different URL. On May 2nd 2020, the questionnaire was augmented - in light of the Covid-19 pandemic - with an extended mood scale, and an instrument comprising 47 items quantifying self-perceived effects on mood, behaviour and outlook (Pandemic General Impact Scale PD-GIS-11). Questions regarding pre-existing psychiatric and neurological conditions, lockdown context, having the virus, and free text fields were added. This coincided with further promotion via BBC2 Horizon and BBC Homepage.

### Data processing and statistical analysis

Analyses were conducted in MATLAB R2020a. Participants with missing data were retained as some questions were contingent on others; therefore, observations per analysis vary with data availability. The questionnaire was organised into the following scales: demographics/lifestyle, online technology use, mood, personality, compulsive traits, and pandemic impact. Where appropriate, scales were summarised in the following steps. Agree-disagree and frequency items were filtered for missing data casewise within scale, converted to numeric, rank transformed to normality, and subscale scores estimated using principal component analysis. Components with eigenvalues>1 were varimax rotated and component scores estimated by regression. PCAs are reported in **Supplement 2**.

Cohort demographics were segmented into Pre-Pandemic, Early-Pandemic, and Mid-Pandemic epochs (**Figure 1a)**. General linear modelling tested how sociodemographic variables statistically predicted differences in mood, anxiety, and sleep, between the densely sampled and demographically matched Pre-Pandemic and Mid-Pandemic epochs.

Subscales of the pandemic impact instrument (PD-GIS-11 **Supplement 3**) quantified self-perceived impact across 11 psycho-socio-economic dimensions. These were cross-validated against the mental health self-assessment using CCA and Pearson’s correlation. General linear modelling (GLM) determined the relationship of sociodemographic variables, home context, cohabitees and work arrangements to the PD-GIS-11. Further GLMs examined carers and people reporting psychiatric and neurologic diagnoses (for groups with N>90 members) with the sociodemographics factored out. Due to the expected shared variance between online technology use, personality traits and compulsivity, their multivariate relationships with the PD-GIS-11 were quantified via canonical correlation analysis (CCA).

Latent Dirichlet Allocation (LDA)^7^ was applied to extracted common topics from free text responses to the questions “What has been most POSITIVE about the lockdown?”, “What has been most NEGATIVE about the lockdown?” and “What have you done that you would recommend to others because it has helped you during the lockdown?” Further details are given in **Supplement 4**. Optimal numbers of topics were estimated for each question as follows. LDA models of different complexity were trained on 2/3rds of the participants’ data. The trained models were applied to the remaining data. The lowest resultant perplexity value, quantifying fit of theoretical and observed topic word distributions, was identified and the LDA model retrained on all data at the corresponding level of complexity. Inter-individual differences in topic mixture estimates were estimated and analysed with reference to mood, PD-GIS-11 and at-risk sub-populations using CCA and t-tests.

## Data Availability

Summary data for this cohort are provided in the supplementary appendices.

## Acknowledgements

Dr Hampshire is supported by the UK Dementia Research Institute and Biomedical Research Centre at Imperial College London with technology development supported by EU-CIG EC Marie-Curie CIG and NIHR grant II-LB-0715-20006. Dr Soreq’s role was supported by MRC grant MR/R005370/1. William Trender is supported by the EPSRC Center for Doctoral Training in Neurotechnology. Dr Chamberlain’s role in this study was funded by a Wellcome Trust Clinical Fellowship (Reference 110049/Z/15/Z). Mitul Mehta is in part supported by the National Institute for Health Research (NIHR) Biomedical Research Centre at South London and Maudsley NHS Foundation Trust and King’s College London. The views expressed are those of the author(s) and not necessarily those of the NHS, the NIHR or the Department of Health and Social Care. We would like to acknowledge COST Action CA16207 ‘European Network for Problematic Usage of the Internet’, supported by COST (European Cooperation in Science and Technology); and the support of the National UK Research Network for Behavioural Addictions (NUK-BA).

## Declaration of interests

Dr. Chamberlain consults for Promentis, and receives stipends for journal editorial work from Elsevier. Dr. Grant has received research grants from the TLC Foundation for Body-Focused Repetitive Behaviors, Biohaven, Promentis, and Avanir Pharmaceuticals. Prof. Mehta has received grant income from Takeda Pharmaceuticals, Johnson & Johnson and Lundbeck. Dr Adam Hampshire is owner and founder of Future Cognition Ltd and H2 Cognitive Designs Ltd, which develop custom cognitive assessment software for other university-based research groups. Dr Peter Hellyer is owner and co-founder of H2 Cognitive Designs Ltd. The authors report no other conflicts of interest.

## Supplementary materials: Hampshire et al. Contents

**Supplement 1 - Study Questionnaires**

**Supplement 2 - Principal Component Analyses of Questionnaire Instrument scales**

**Supplement 3a - PD-GIS individual item and PCA**

**Supplement 3b - Canonical correlation analysis between Mood State and PD-GIS-11**

**Supplement 3c - Demographic predictors of the PD-GIS-11**

**Supplement 3d - Rates of previously diagnosed clinical conditions reported in the sample**

**Supplement 3e - GLM for clinical conditions vs PD-GIS-11 subscales**

**Supplement 3f - sub-sampling and permutation analyses to characterise overfit and reproducibility of canonical correlation analysis of PD-GIS-11 items vs traits and technology use**

**Supplement 4a - topics and their top ten exemplars**

**Supplement 4b - CCA of free text topic estimates vs mood self assessment item scores**

**Supplement 4c - CCA of free text topic estimates vs items of the PD-GIS-11**

**Supplement 4d - Topics, their relative prevalence in select sub-populations and top ten exemplars selected as having the highest topic mixtures**

## Supplement 1 - Study Questionnaires

### (a) Demographic and other contextual information

The following background information was collected from participants: age, gender, ethnicity, country of residence, level of education, occupational status, and income.

### (b) Mood, anxiety, and sleep

Mood and anxiety symptoms were recorded using the extensively validated Patient Health Questionnaire 2 (PHQ-2) and GAD-7 respectively ^1,2^The PHQ-2 and GAD-7 ask about symptoms over the preceding two weeks, and each question is answered on a 4-point scale, from 0 (not at all) to 3 (nearly every day). Additionally, we asked how many hours on average participants slept per night.

### (c) Personality traits, and compulsivity

Personality traits were quantified using the extensively validated Big-5 Inventory, which comprises 44 questions^3^. Each question is a short phrase and is answered on a 5-point rating scale from 1 (strongly disagree) to 5 (strongly agree). Aspects of personality classically reflect extraversion, agreeableness, conscientiousness, neuroticism and openness to experience ^3^Based on prior factor analysis of data from 60,000 participants, we used an abbreviated version, comprising 18 questions with a data-driven structure of 6 components. These are reported in the factor analysis in **Appendix 2**.

Compulsivity is a trans-diagnostic concept representing the tendency towards repetitive habits, and was measured using the Cambridge-Chicago Compulsivity Trait Scale (CHI-T)^4^. This is a 15-item questionnaire that is answered on a 4-point rating scale ranging from 1 (strongly disagree) to 4 (strongly agree). The CHI-T is sensitive to compulsivity across a range of disorders ^4,5^.

### (d) Impact of the pandemic

The Pandemic General Impact Scale (PD-GIS-11) was developed for the current study to quantify the self-reported negative and positive impacts of the Covid-19 pandemic, on multiple levels of psycho-socio-economic investigation. The instrument comprised 47 questions, relating to potential negative and positive aspects of the situation, and longer term outlook. Each item is answered on a 5-point scale ranging from 1 (strongly disagree) to 4 (strongly agree). Negative impact questions covered areas of concern for health (own health and that of others), being concerned with the consequences of contracting Covid-19, loneliness, conflict at home, negative emotions from reading/listening to news, grieving, loss of employment/job/income, loss of leisure and well being activities, loss of daily structure, disruption of sleep patterns, less healthy lifestyles, less focus on personal hygiene, loss of productivity, social disconnection, life being dominated by infection control routines, loss of important goods/medication/services, more arguments in the household, and going on the internet to avoid people at home. Positive impact questions covered less commuting time, more structure to the day, joy at being able to spent more time with people at home, more connections with people online, sense of shared community, more efficient/productive work, being more relaxed due to more time at home, better sleep due to spare time, greater sense of purpose in work, greater opportunity to exercise, improved natural environment, time to read for pleasure, work less stressful due to doing it from home, spending more time on hobbies, spending less and saving money, more social contact outside of the home, feeling less tired, feeling better connected with people at home, more wildlife, taking greater appreciation for the simple things in life, and being less stressed by daily responsibilities.

### (e) Online Technology use

Technology use was quantified by asking about frequency of use of the following, over the previous 4 week period: Smart Phone, Computer (Desktop/Laptop), Tablet Device, Gaming Console, Email, Social Media, reading the news, playing computer games, online gambling, working, learning/studying, shopping, streaming films or music, and searching for information online. Each question was responded to on a 7-point scale, from 0 (never) to 7 (more often than hourly every day).

### (f) Stress from online technology

Stress from online technology was measured by asking the participants the following questions, regarding the past 4 weeks: When you checked Email, did it tend to make you feel stressed/unhappy or relieved/happy? When you used social media, did it tend to make you feel stressed/unhappy or relieved/happy? When you read the news, did it tend to make you feel stressed/unhappy or relieved/happy? When you played computer games, did it tend to make you feel stressed/unhappy or relieved/happy? The response options for each question were: “Mostly stressed/unhappy”,”Mostly relieved/happy”,”Both”, or “Neither”.

### (g) Maladaptive (‘Addictive’) use of online technology

Maladaptive use of online technology was quantified using the following questions, which were based on expert consensus amongst the study team in the field of Problematic Usage of the Internet: How often did you check email or social media accounts after you went to bed? How often did you use internet related activities to block out disturbing thoughts or soothe yourself? How often did you choose to spend time on internet related activities to battle loneliness or boredom? How often did you suffer from negative financial consequences because of an online activity? How often did you check your email or social media account or equivalent before something else that you needed to do? How often did you try to stop an excessive online activity but feel a compulsion to continue? How often did you try to cut down the amount of time you spend on-line and fail? The questions asked about these areas over the preceding 4 week period. For the first question (using technology before bed), response options were 1 (never) to 5 (daily). For the other questions, response options were: 1 (never) to 7 (more than hourly every day).

---

**Appendix 2a.**
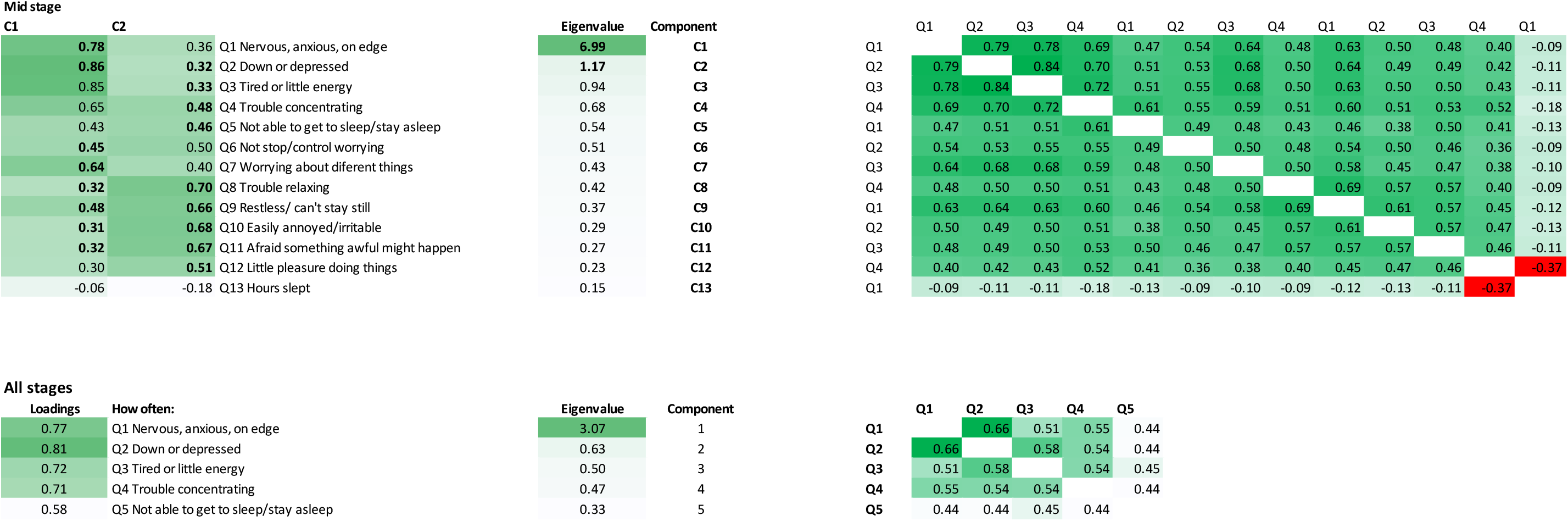
PCA of the Mood Self Assessment.

**Appendix 2b.**
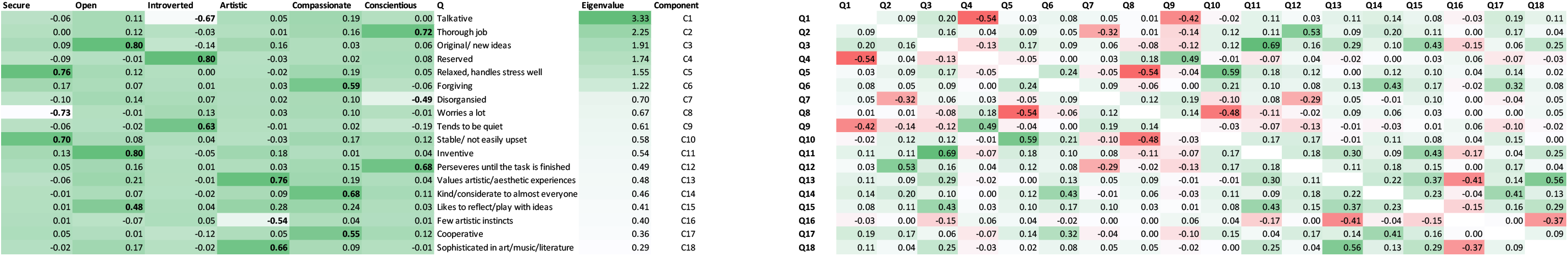
PCA of the Minimise Big5 Personality Scale.

**Appendix 2c.**
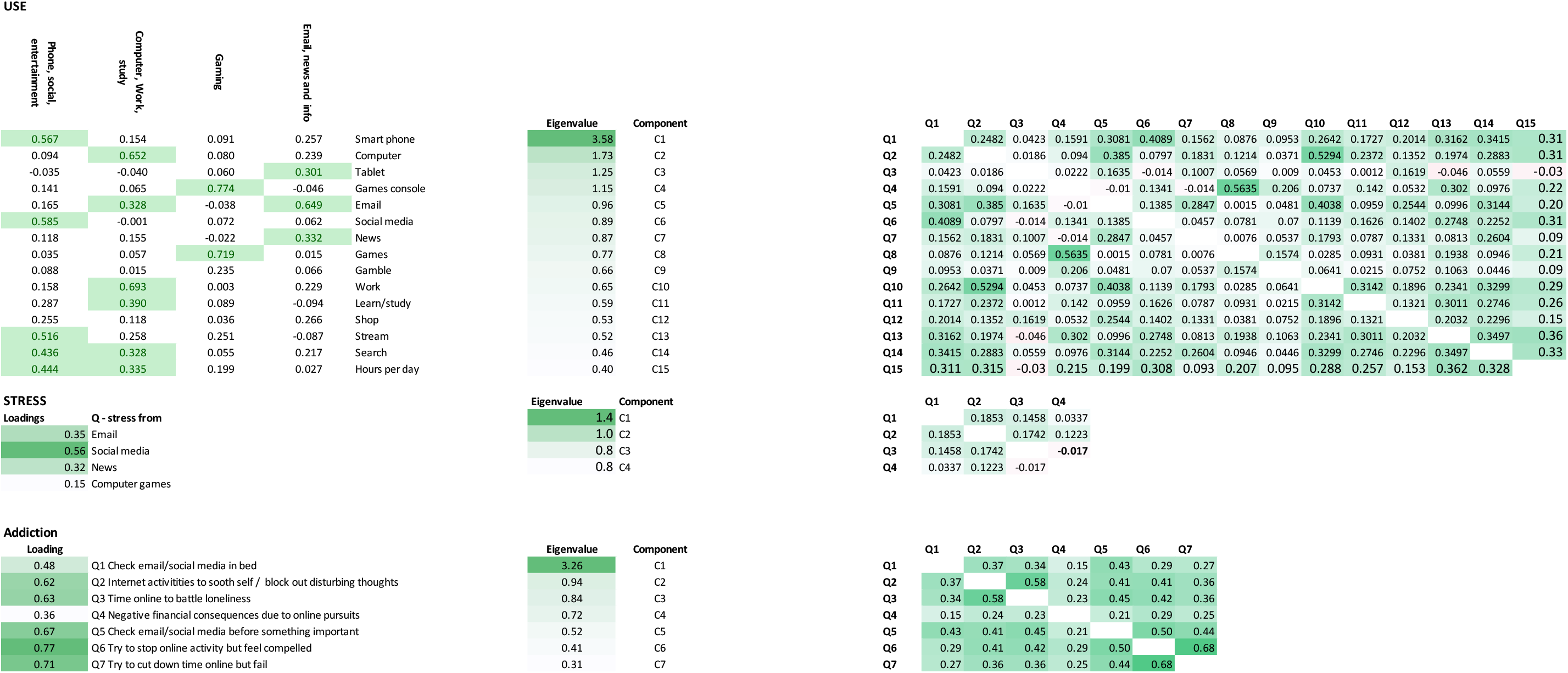
PCA of the Technology use, stress and addiction scales.

**Appendix 2d.**
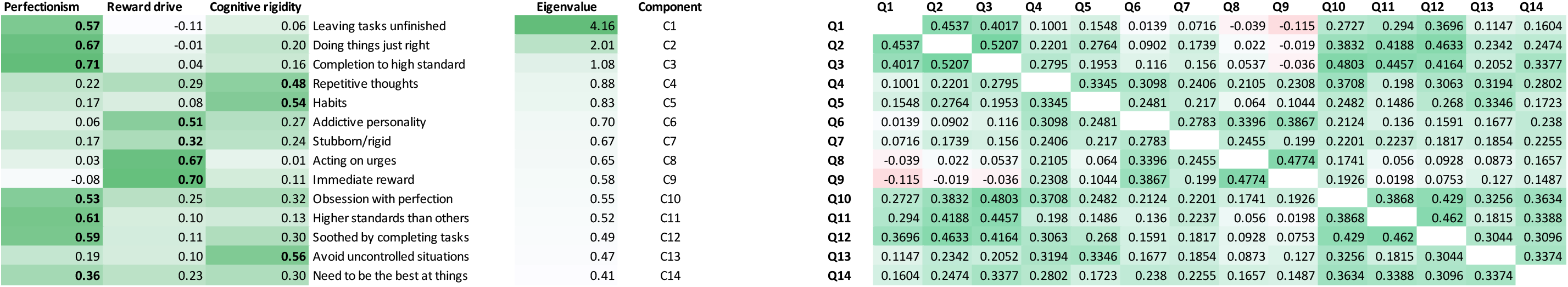
PCA of the CHIT compulsivity scale.

**Appendix 3a.**
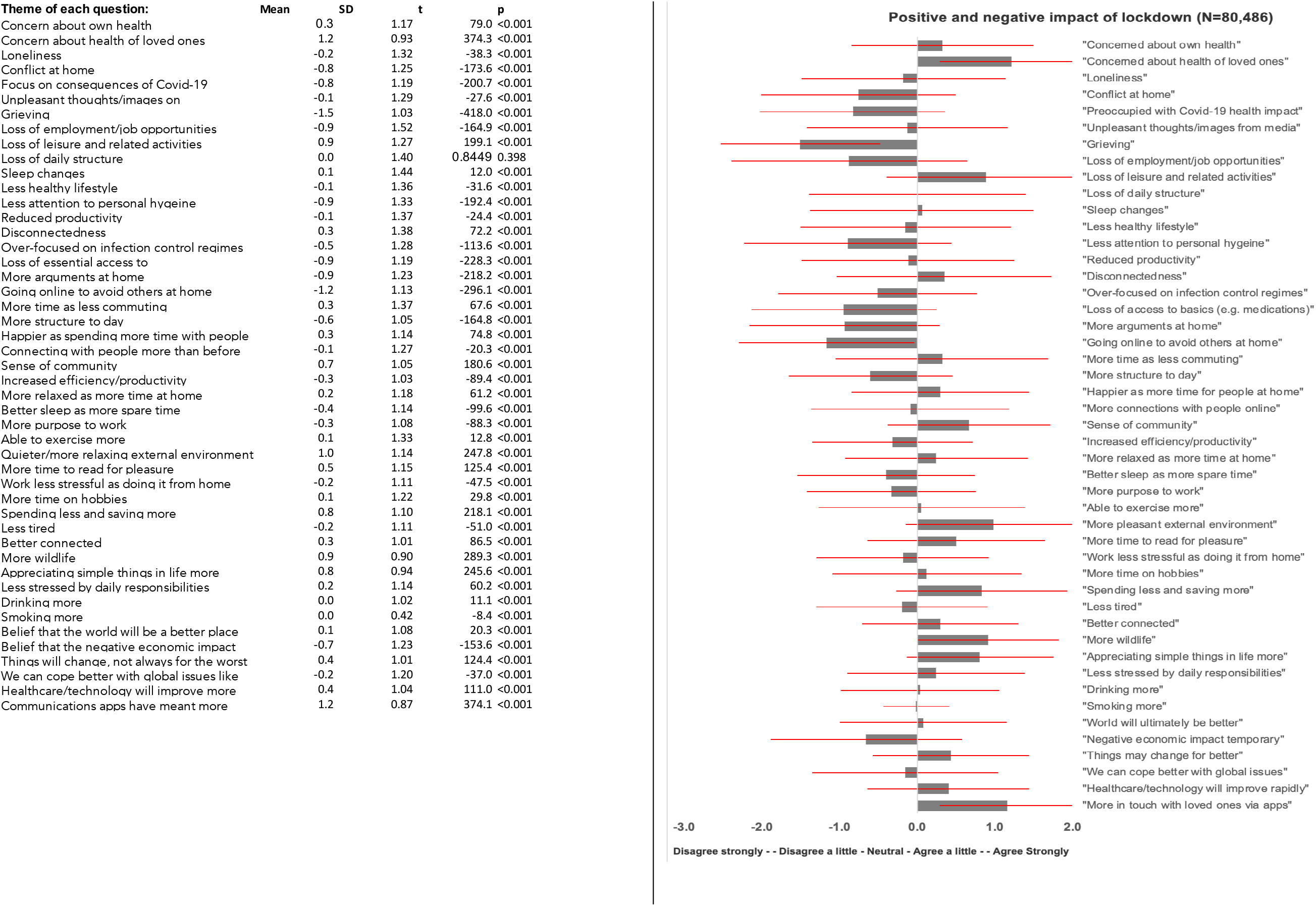
PD-GIS-11 response distributions.

**Appendix 3b.**
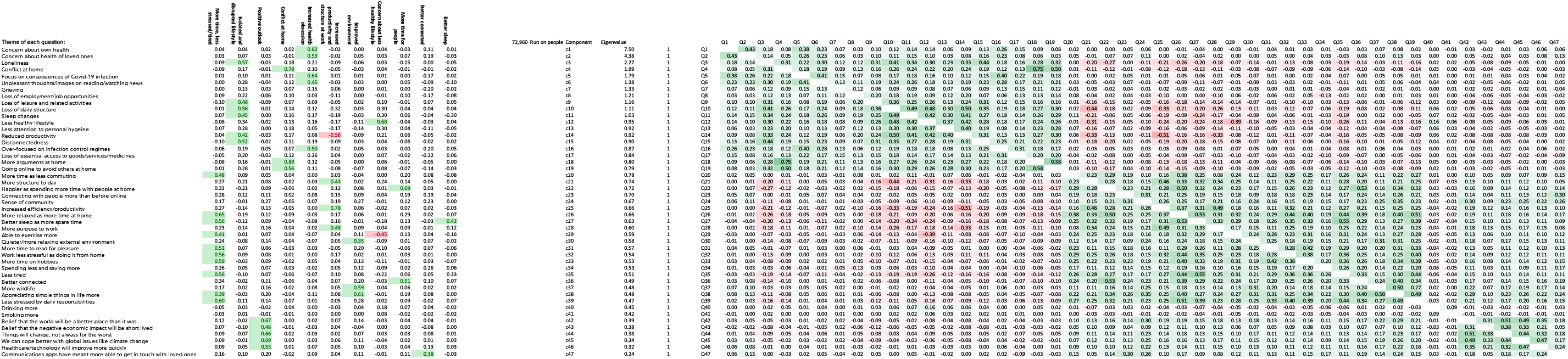
PCA of PD-GIS-11 (zoom to read)

**Appendix 3c.**
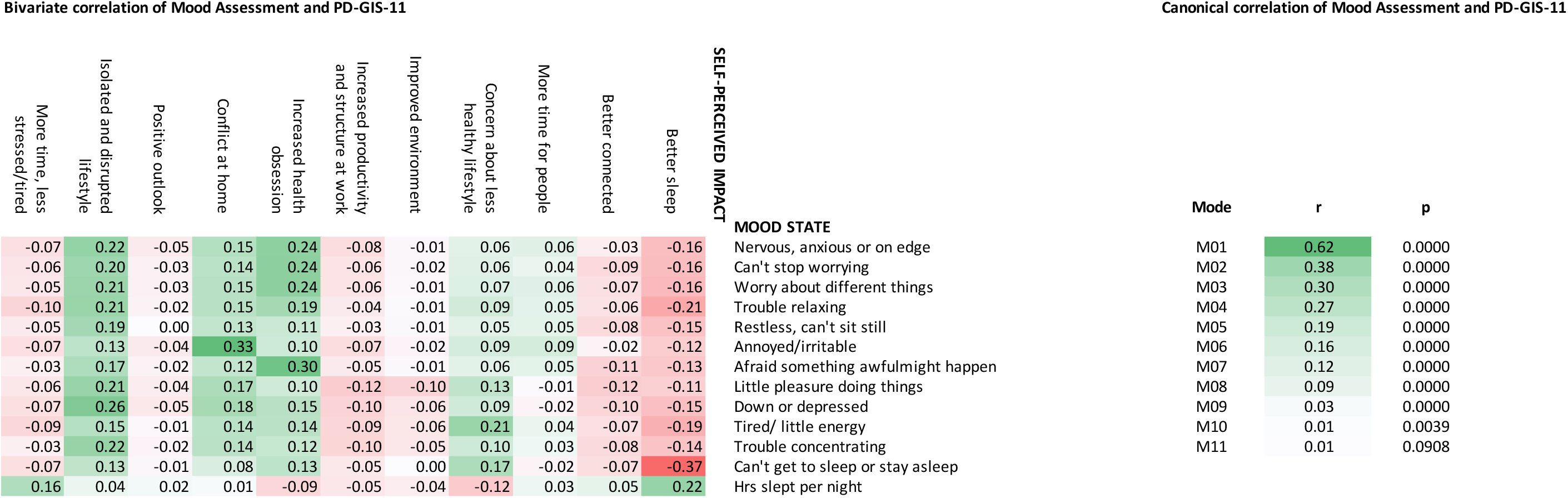
PD-GIS-11 vs Mood Self Assessment.

**Appendix 3d.**
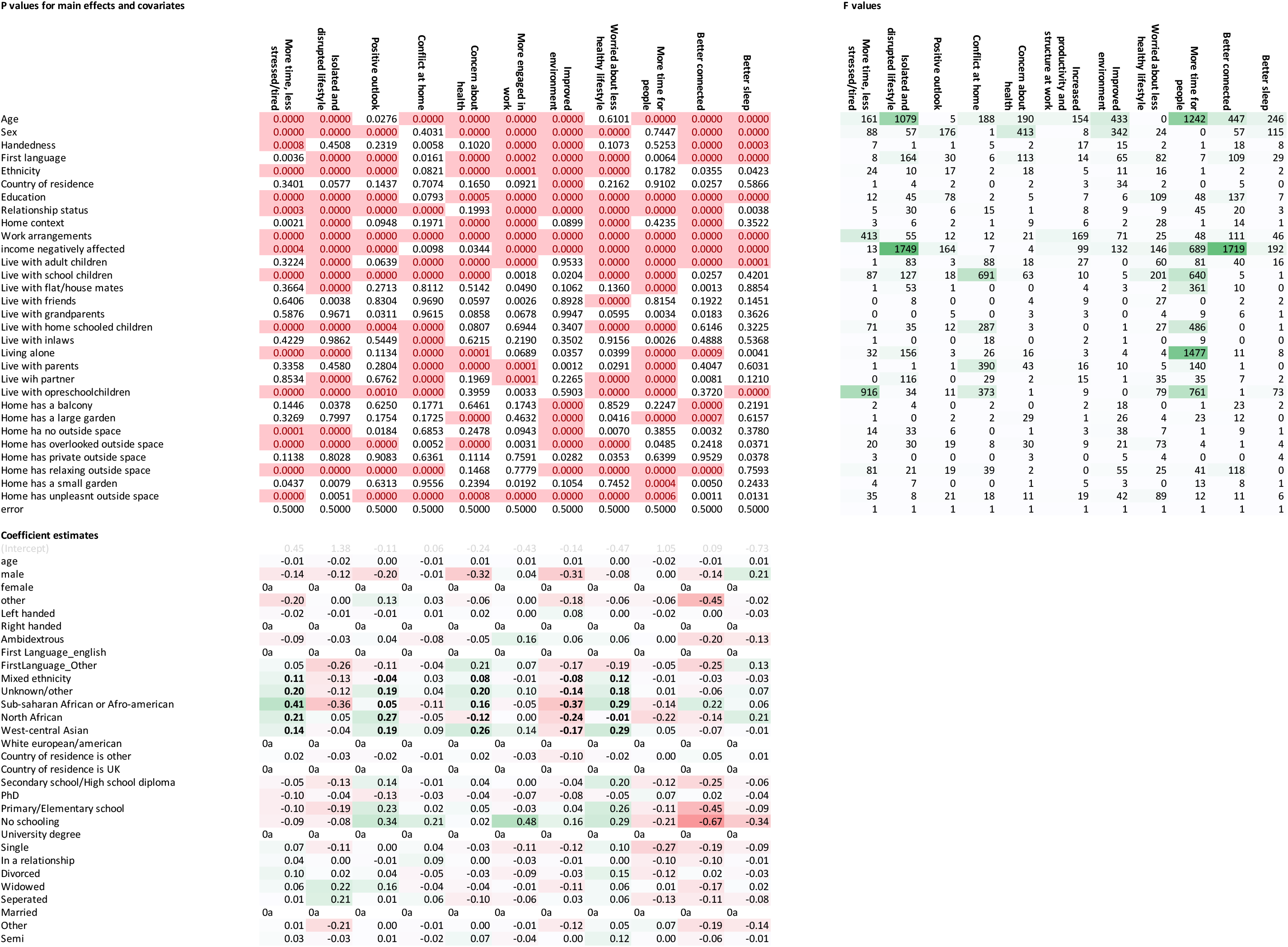
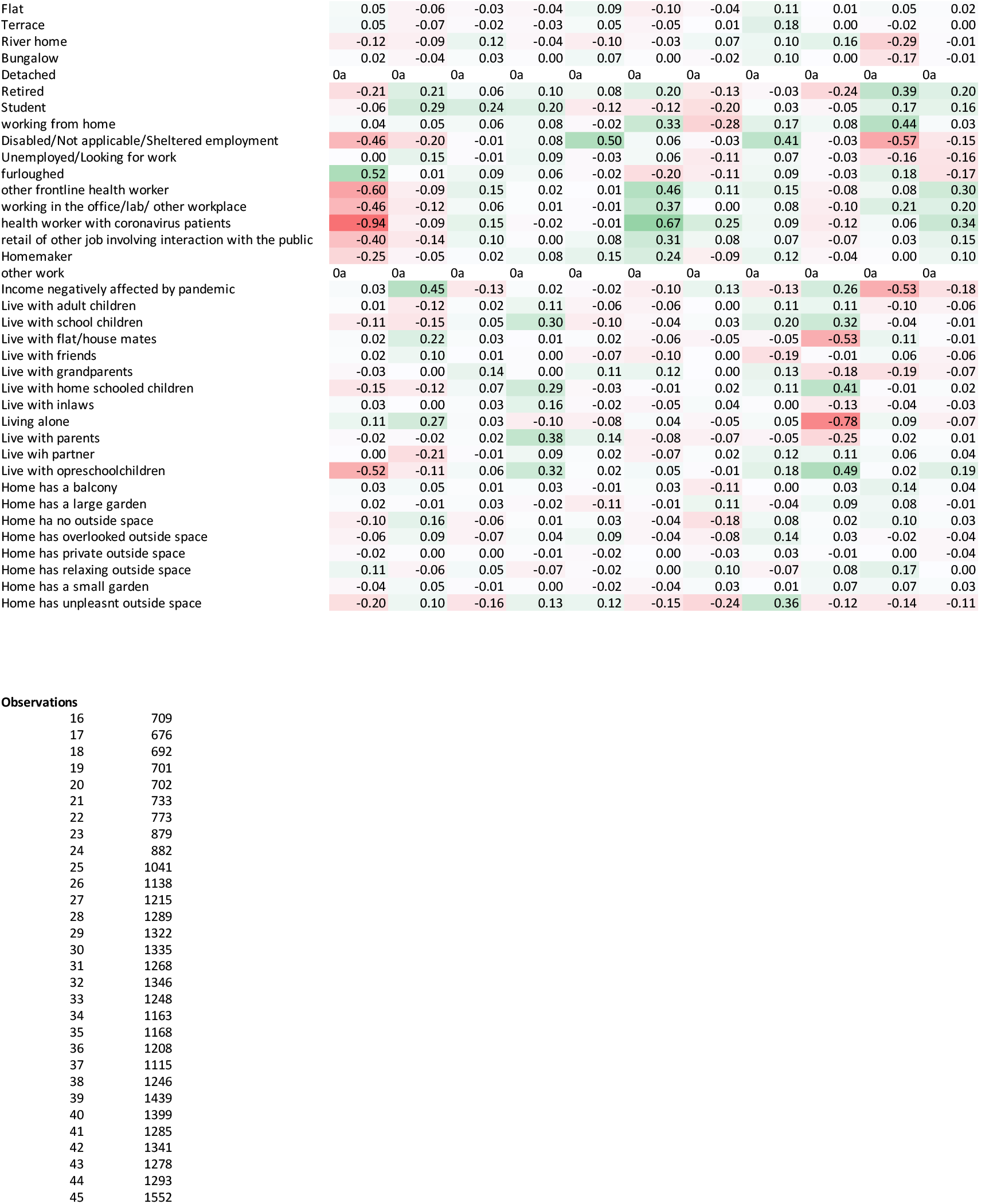
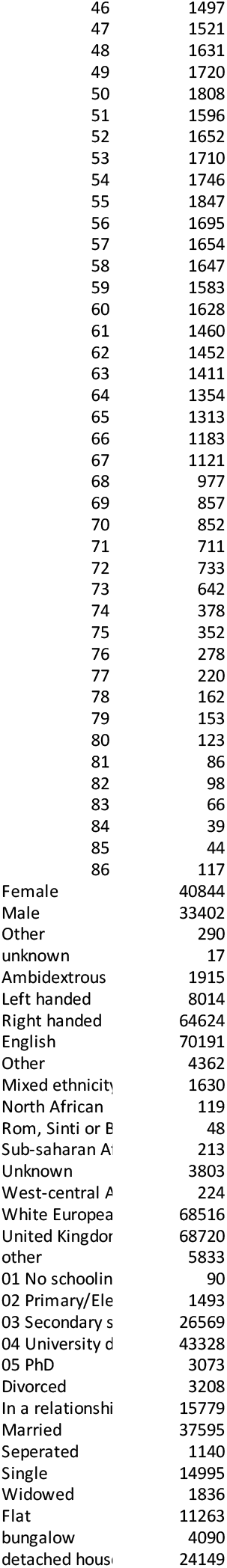
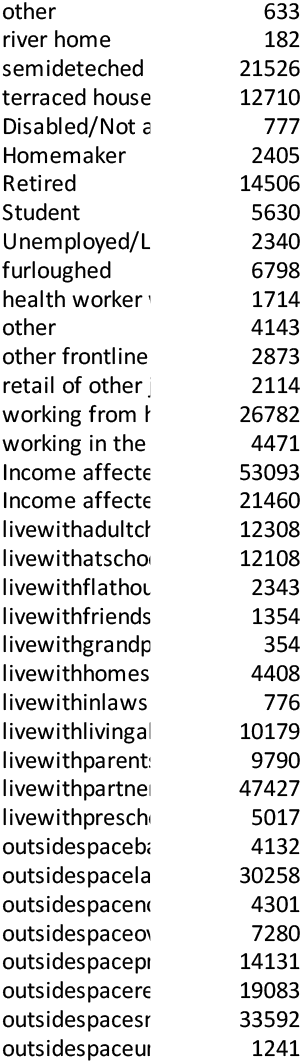
PD-GIS-11 vs socio-demographic, economic and context variables.

**Appendix 3e.**
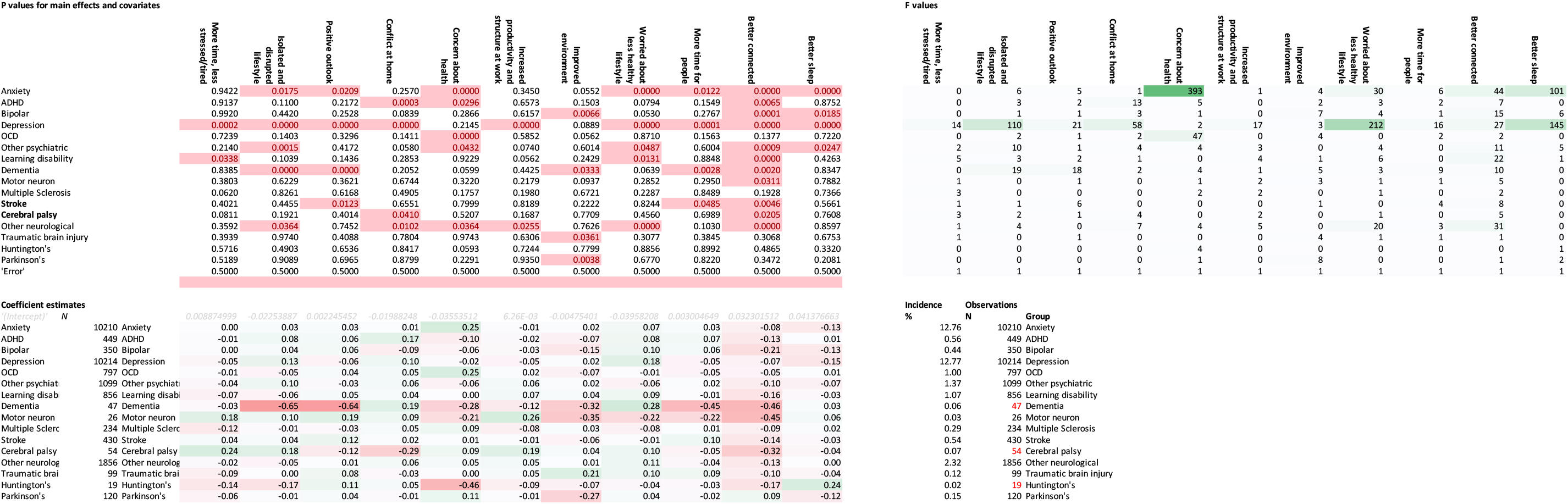
PD-GIS-11 vs preexisting conditions.

**Appendix 3f.**
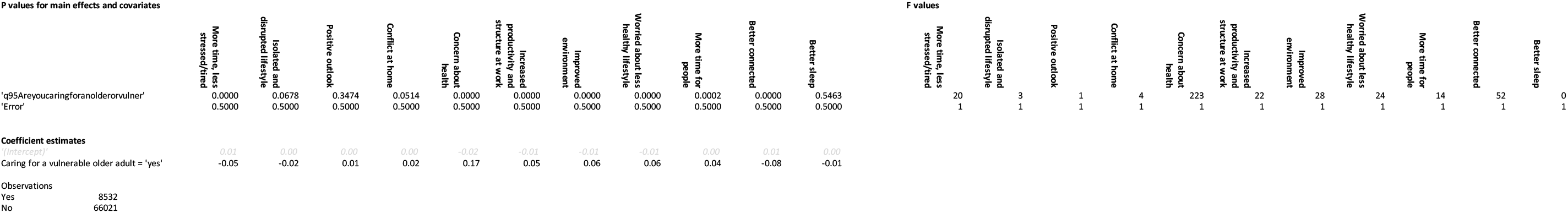
PD-GIS-11 vs caring for older adult.

**Appendix 3g.**
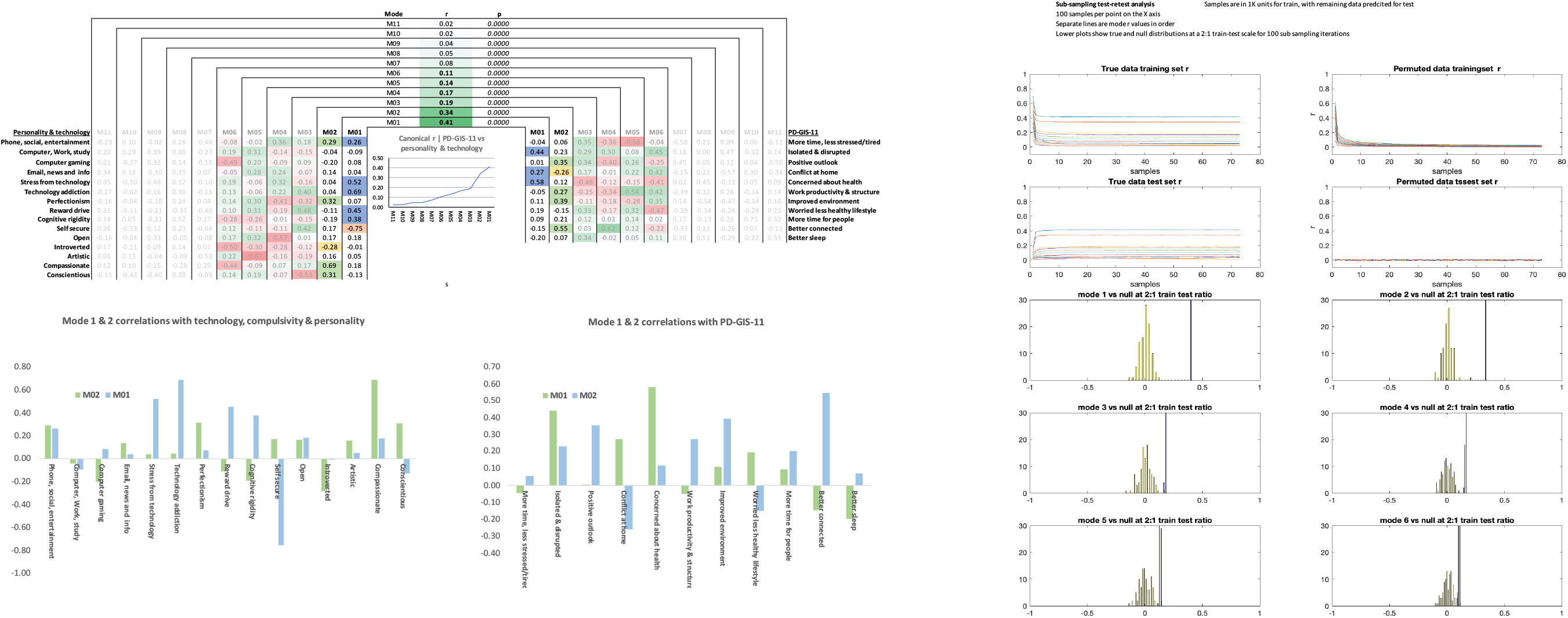
PD-GIS-11 vs traits and technology variables.

**Appendix 5a.**
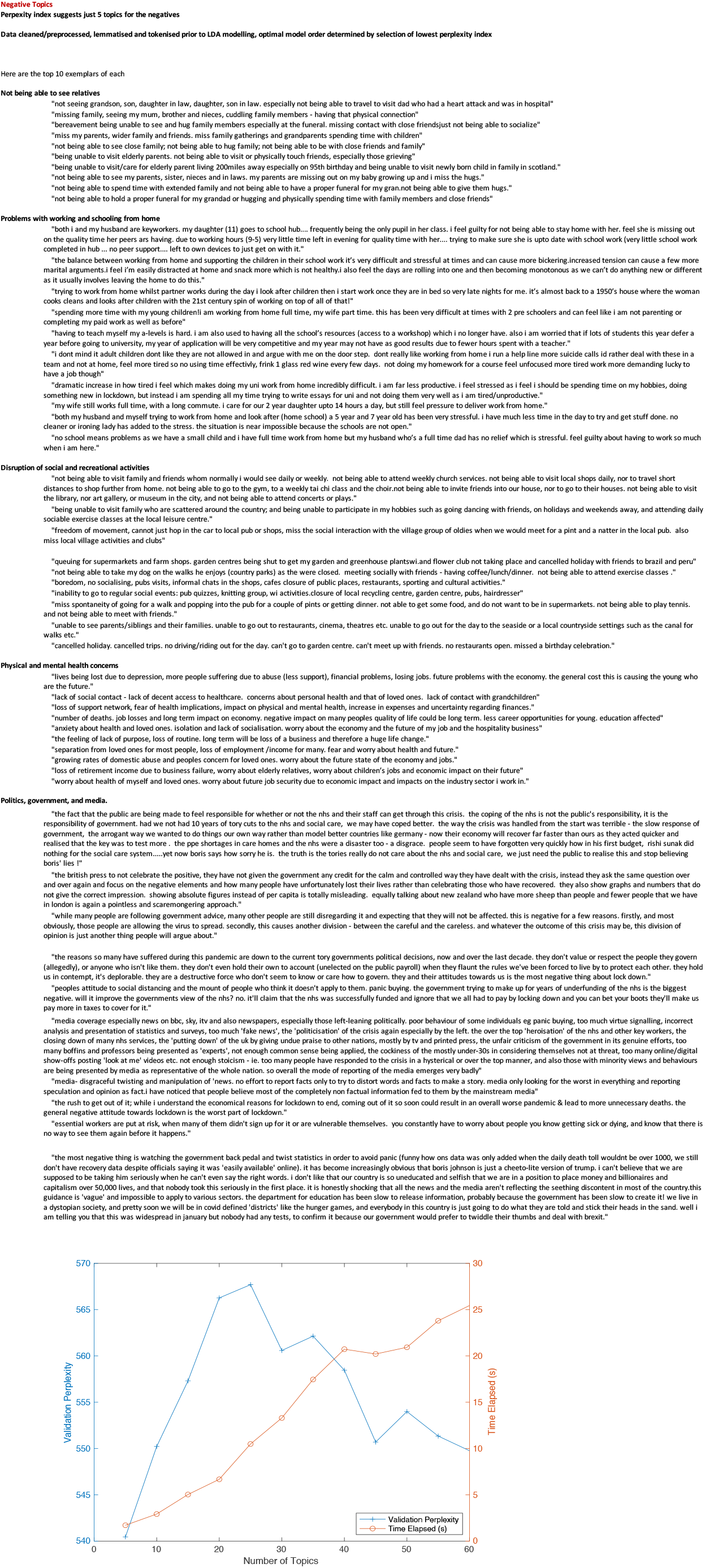
negative topics (zoom to read)

**Appendix 5b.**
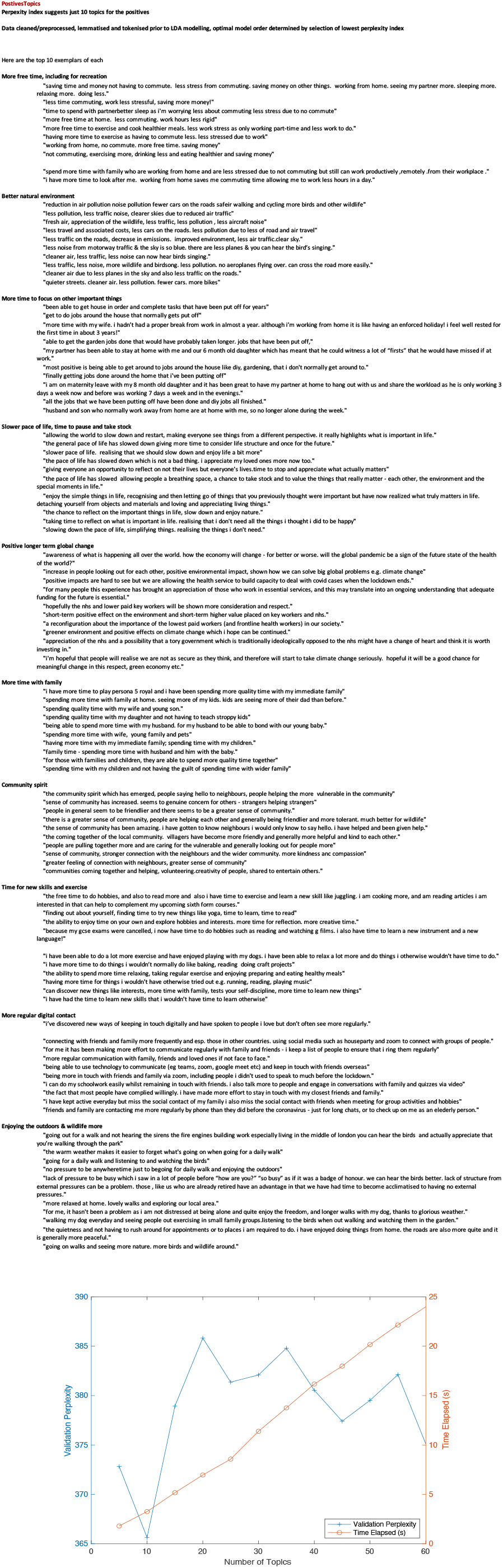
positive topics (zoom to read)

**Appendix 4c.**
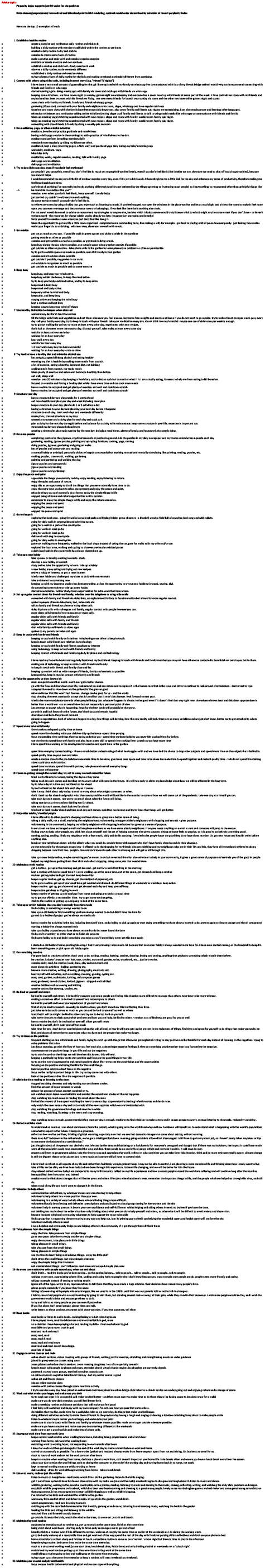
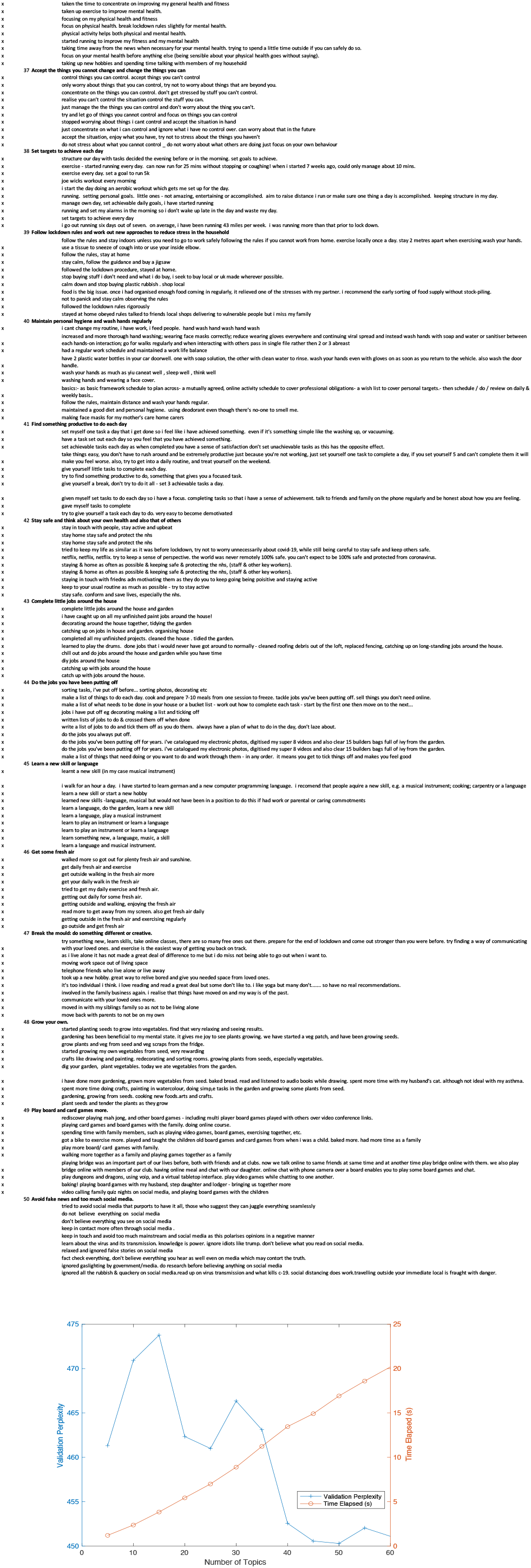
Advice topics (zoom to read)

**Appendix 4d.**
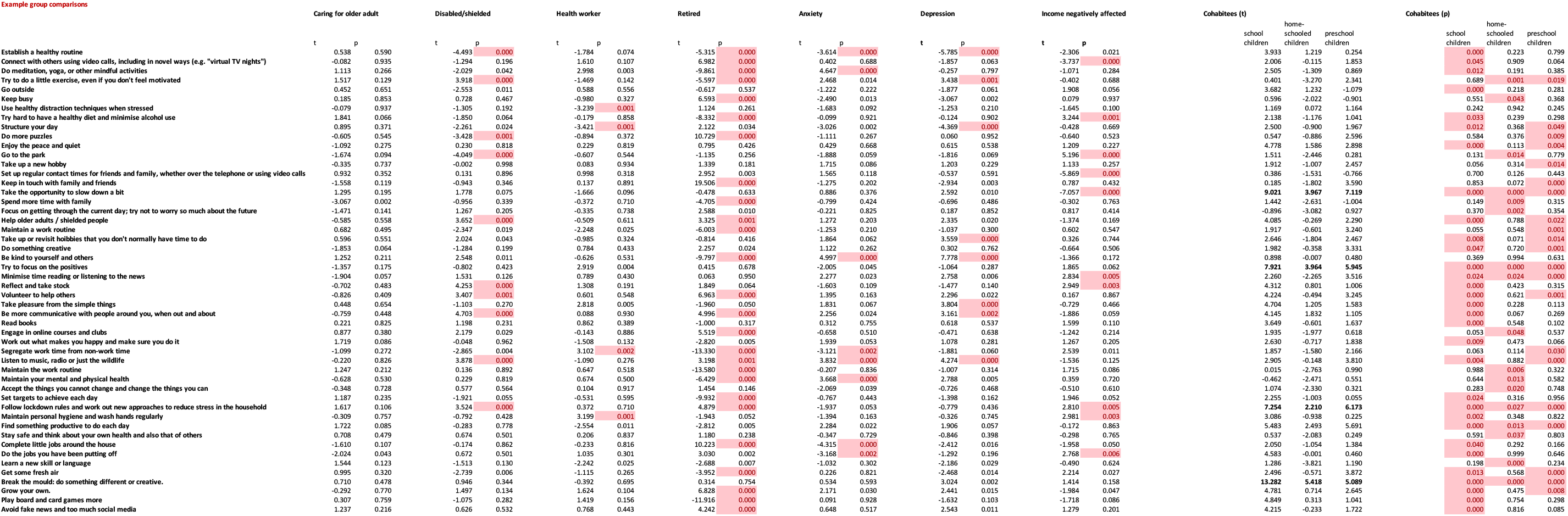
Advice topic prevalences vs risk factors (zoom to read)

**Appendix 4e.**
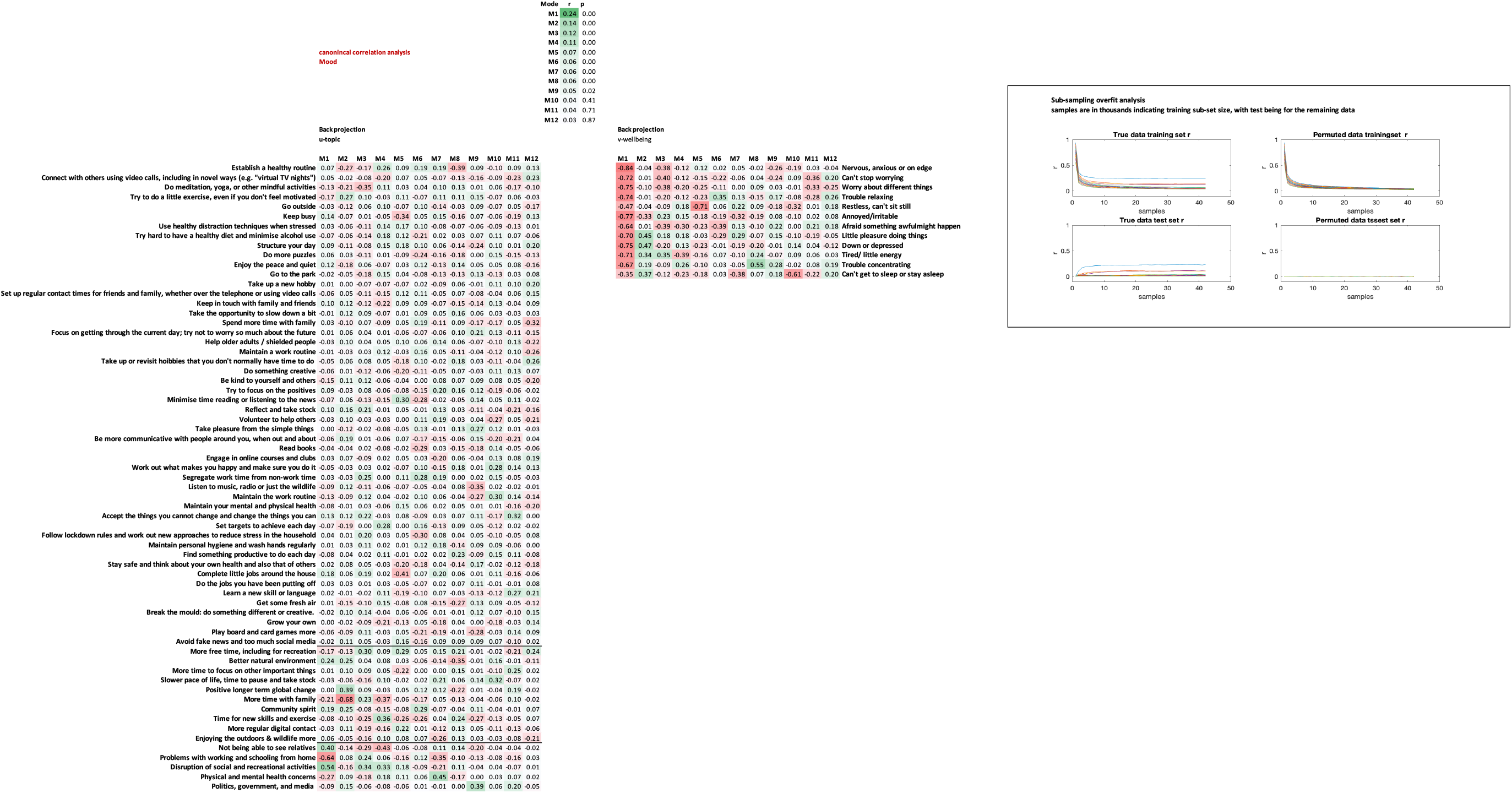
CCA of all topics vs mood self assessment (zoom to read)

**Appendix 4f.**
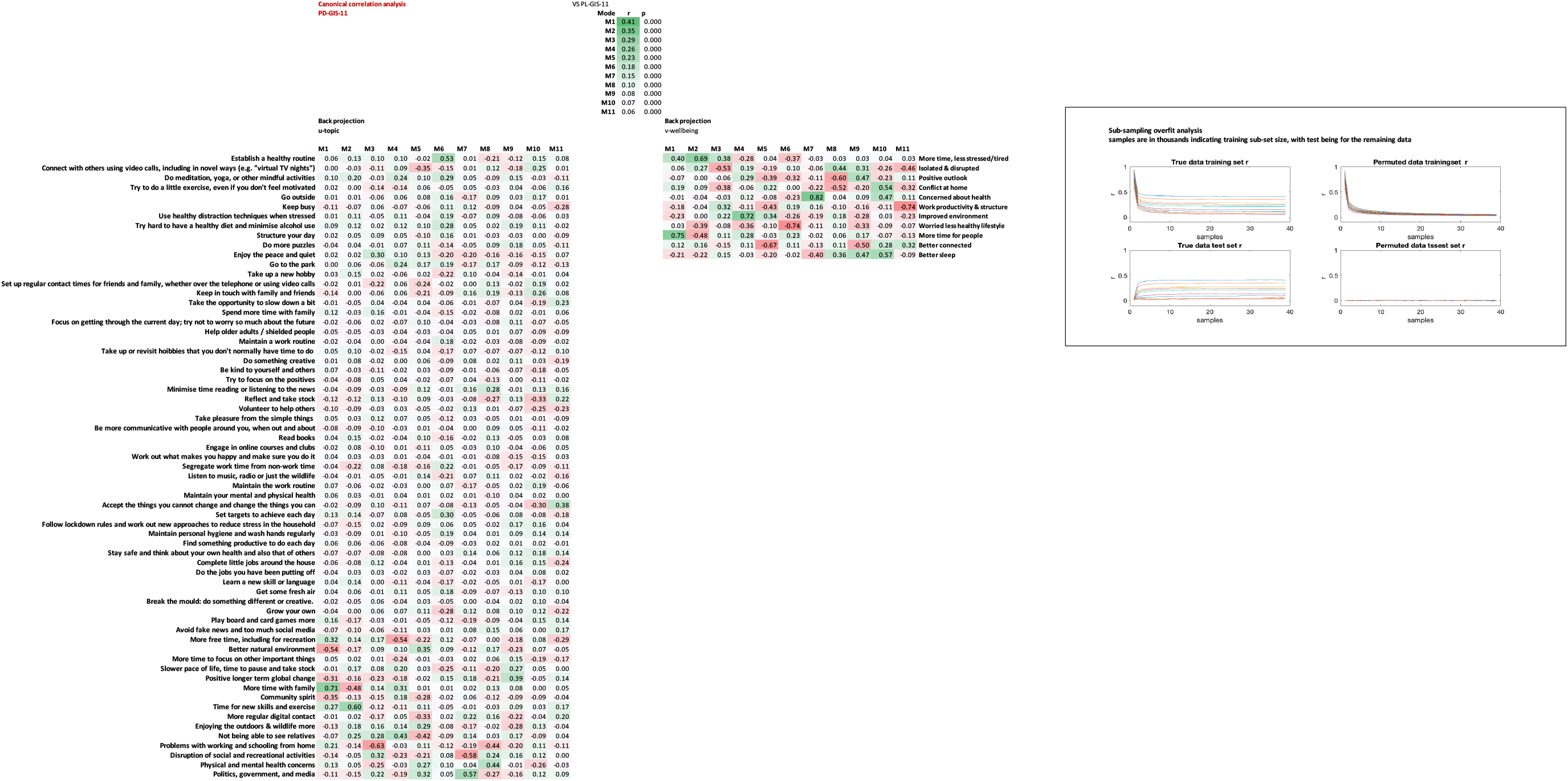
CCA of all topics vs PL-GIS-11 (zoom to read)

## Notes

### Competing Interest Statement

Disclosures: Dr. Chamberlain consults for Promentis, and receives stipends for journal editorial work from Elsevier. Dr. Grant has received research grants from the TLC Foundation for Body-Focused Repetitive Behaviors, Biohaven, Promentis, and Avanir Pharmaceuticals. Prof. Mehta has received grant income from Takeda Pharmaceuticals, Johnson & Johnson and Lundbeck.

### Clinical Trial

This was a large scale observational study run in collaboration with BBC2 Horizon. It was not a clinical trial.

### Author Declarations

Imperial College Research Ethics Committee. 17IC4009 ICREC

### Summary of Updates

Extended analyses have been added in this revision.

## References

1 Pfefferbaum, B. & North, C. S. Mental Health and the Covid-19 Pandemic. N Engl J Med, doi:10.1056/NEJMp2008017 (2020).

2 Holmes, E. A. et al. Multidisciplinary research priorities for the COVID-19 pandemic: a call for action for mental health science. Lancet Psychiatry 7, 547–560, doi:10.1016/S2215-0366(20)30168-1 (2020).

3 Kiraly, O. et al. Preventing problematic internet use during the COVID-19 pandemic: Consensus guidance. Compr Psychiatry 100, 152180, doi:10.1016/j.comppsych.2020.152180 (2020).

4 Pareek, M. et al. Ethnicity and COVID-19: an urgent public health research priority. Lancet 395, 1421–1422, doi:10.1016/S0140-6736(20)30922-3 (2020).

5 Brooks, S. K. et al. The psychological impact of quarantine and how to reduce it: rapid review of the evidence. Lancet 395, 912–920, doi:10.1016/S0140-6736(20)30460-8 (2020).

6 Ioannidis, K., Askelund, A. D., Kievit, R. A. & van Harmelen, A. L. The complex neurobiology of resilient functioning after childhood maltreatment. BMC Med 18, 32, doi:10.1186/s12916-020-1490-7 (2020).

7 Blei D.M N. A. Y., Jordan M.I, Lafferty J. Latent Dirichlet allocation. Journal of Machine Learning Research 3, 993–1022 (2003).

8 Chowkwanyun, M. & Reed, A. L.,Jr. Racial Health Disparities and Covid-19 - Caution and Context. N Engl J Med, doi:10.1056/NEJMp2012910 (2020).

9 England., P. H. Disparities in the risk and outcomes of COVID-19‥ (2020).

10 Kirby, T. Evidence mounts on the disproportionate effect of COVID-19 on ethnic minorities. Lancet Respir Med 8, 547–548, doi:10.1016/S2213-2600(20)30228-9 (2020).

11 Lai, J. et al. Factors Associated With Mental Health Outcomes Among Health Care Workers Exposed to Coronavirus Disease 2019. JAMA Netw Open 3, e203976, doi:10.1001/jamanetworkopen.2020.3976 (2020).

12 Rossi, R. et al. Mental Health Outcomes Among Frontline and Second-Line Health Care Workers During the Coronavirus Disease 2019 (COVID-19) Pandemic in Italy. JAMA Netw Open 3, e2010185, doi:10.1001/jamanetworkopen.2020.10185 (2020).

13 Chamberlain, S. R., Stochl, J., Redden, S. A. & Grant, J. E. Latent traits of impulsivity and compulsivity: toward dimensional psychiatry. Psychol Med 48, 810–821, doi:10.1017/S0033291717002185 (2018).

14 Cortese, S. & Coghill, D. Twenty years of research on attention-deficit/hyperactivity disorder (ADHD): looking back, looking forward. Evid Based Ment Health 21, 173–176, doi:10.1136/ebmental-2018-300050 (2018).

15 Posner, J., Polanczyk, G. V. & Sonuga-Barke, E. Attention-deficit hyperactivity disorder. Lancet 395, 450–462, doi:10.1016/S0140-6736(19)33004-1 (2020).

16 Grant J.E P. A., Chamberlain S.R.. Obsessive-Compulsive Personality Disorder. American Psychiatric Association Publishing (2019).

17 Ioannidis, K. et al. Problematic internet use (PIU): Associations with the impulsive- compulsive spectrum. An application of machine learning in psychiatry. J Psychiatr Res 83, 94–102, doi:10.1016/j.jpsychires.2016.08.010 (2016).

18 Tiego, J. et al. Problematic use of the Internet is a unidimensional quasi-trait with impulsive and compulsive subtypes. BMC Psychiatry 19, 348, doi:10.1186/s12888-019-2352-8 (2019).

19 Fancourt D S. A., Bu F. Trajectories of depression and anxiety during enforced isolation due to COVID-19; longitudinal analyses of 59,318 adults in the UK with and without diagnosed mental illness. medRxiv (2020).

20 Wang, C. et al. A longitudinal study on the mental health of general population during the COVID-19 epidemic in China. Brain Behav Immun, doi:10.1016/j.bbi.2020.04.028 (2020).

## References

1 Kroenke, K., Spitzer, R.L. & Williams, J.B. The Patient Health Questionnaire-2: validity of a twoitem depression screener. Med Care 41, 1284-1292, doi:10.1097/01.MLR.0000093487.78664.3C (2003).

2 Spitzer, R.L., Kroenke, K., Williams, J.B. & Lowe B.A brief measure for assessing generalized anxiety disorder: the GAD-7. Arch Intern Med 166, 1092-1097, doi:10.1001/archinte.166.10.1092 (2006).

3 John O.P, Naumann L.P & C.J., S. in Handbook of personality: Theory and research (ed R.W. Robins O.P. John, & L.A. Pervin) (Guilford Press, 2008).

4 Chamberlain, S.R. & Grant, J.E. Initial validation of a transdiagnostic compulsivity questionnaire: the Cambridge-Chicago Compulsivity Trait Scale. CNS Spectr 23, 340-346 doi:10.1017/S1092852918000810 (2018).,

5 Albertella, L. et al. Compulsivity is measurable across distinct psychiatric symptom domains and is associated with familial risk and reward-related attentional capture. CNS Spectr, 1-8, doi:10.1017/S1092852919001330 (2019).

